# Solid evidence and liquid gold: trade-offs of processing settled solids, whole influent, or centrifuged influent for co-detecting viral, bacterial, and eukaryotic pathogens in wastewater

**DOI:** 10.1101/2025.03.01.25323156

**Authors:** Sooyeol Kim, Denise Garcia, Caroline McCormack, Rui Xin Tham, Megan E. O’Brien, Erica R. Fuhrmeister, Kara L. Nelson, Rose S. Kantor, Amy J. Pickering

**Affiliations:** Department of Civil and Environmental Engineering, University of California, Berkeley; Department of Environmental and Occupational Health Sciences, University of Washington; Department of Civil and Environmental Engineering, University of Washington; Physical and Life Sciences Directorate, Lawrence Livermore National Laboratory, Livermore, CA; Chan Zuckerberg Biohub San Francisco, San Francisco, California; Blum Center for Developing Economies, University of California

**Author notes:** co-first authors (Sooyeol Kim, Denise Garcia).

**Keywords:** Wastewater surveillance, method comparison, pathogen and antibiotic resistance gene detection, sample matrix comparison

## Abstract

Effective methods for simultaneously measuring viral, bacterial, protozoan, and fungal pathogens in wastewater are needed. Here, we investigate how sample type and nucleic acid extraction protocols affect broad-range pathogen detection. We compared methods for analyzing wastewater solids and whole influent by dPCR detection of spiked and endogenous targets including DNA and RNA viruses (mpox, norovirus, SARS-CoV-2), bacteria (*Clostridium difficile, Campylobacter jejuni*), protozoa (*Cryptosporidium spp*.), fungi (*Candida auris*), and antibiotic resistance genes. Using selected methods, we then analyzed date-matched 1) solid, 2) centrifuged influent, and 3) whole influent samples collected from eleven facilities at three time points. We demonstrate that one workflow can be used to simultaneously detect all targets and that all sample types yielded similar detection levels and nucleic acid concentrations. Comparing normalization of targets by concentration of PMMoV, carjivirus, and 16S, we show that using different controls together can complicate interpretation of concentrations across targets. Centrifuged influent produced comparable or higher target concentrations overall, suggesting that centrifuged influent is a viable option when settled solids are not available and can circumvent the limitation of varying residence times for primary settled solids.

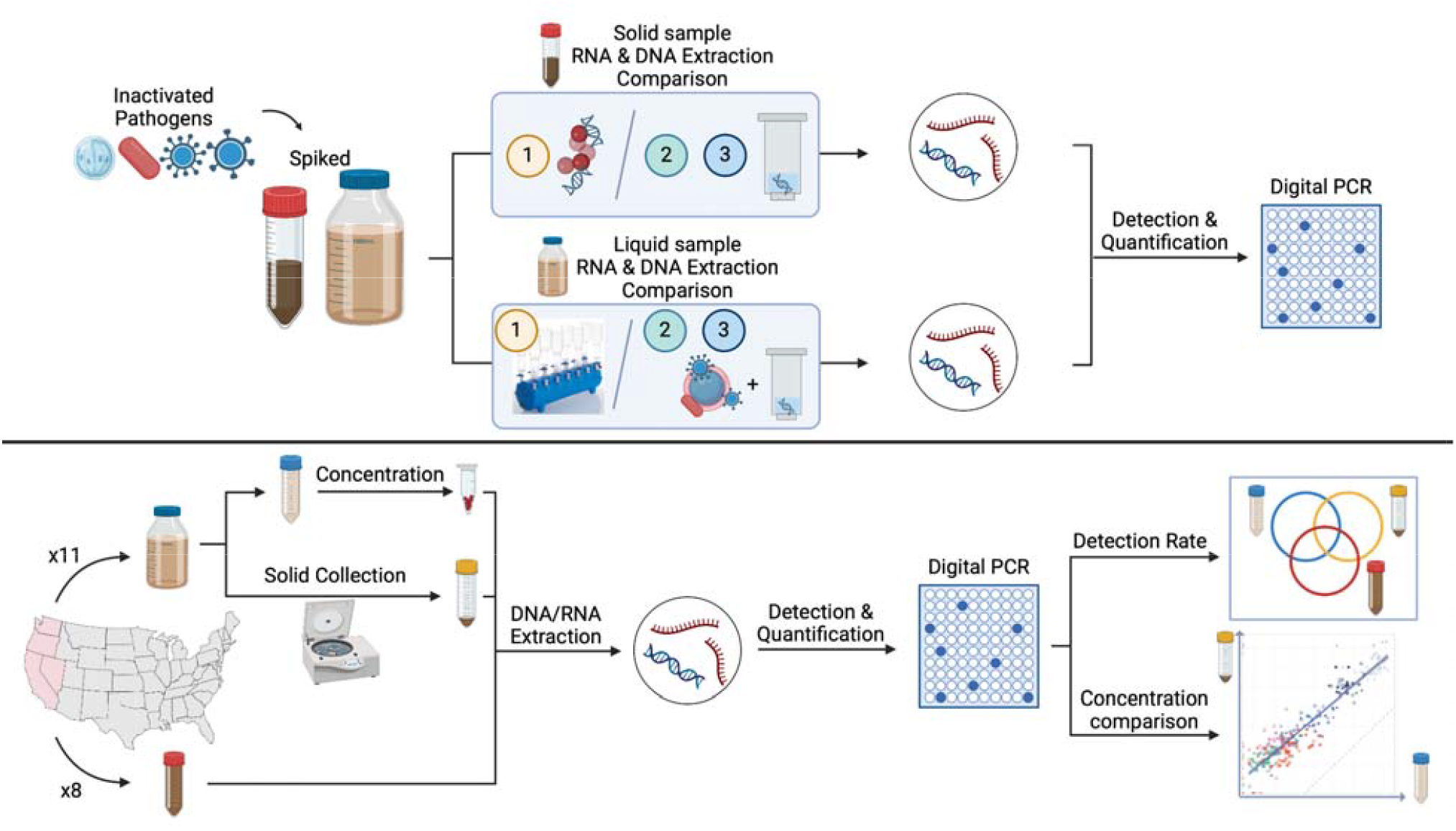

**Synopsis:** We report successful simultaneous detection of viral, bacterial, and eukaryotic pathogens in both solid and liquid wastewater samples and explore multiple ways to compare sample concentrations measured in different matrices.

## 1. Introduction

Wastewater monitoring has emerged as a pivotal tool for assessing population health, offering complementary insights to clinical testing. Wastewater serves as a pooled biological sample from contributing communities and is unaffected by test-seeking behavior^1^, test availability^2^, or reporting delays^3^, which are critical limitations of clinical testing. Successful use of wastewater surveillance during the COVID-19 pandemic has presented an opportunity to utilize the developed infrastructure for broader applications beyond SARS-CoV-2 monitoring. There is an increased interest by the United States Centers for Disease Control and Prevention in expanding wastewater monitoring to include other pathogens of broader public health concern and antibiotic resistance, which could greatly enhance the value of wastewater analysis. Despite significant progress in detecting SARS-CoV-2 and other viruses in wastewater, research on methods for reliable detection of bacterial^4^, protozoan^5^, and fungal^6^ pathogens remains limited. Establishing a standardized technique for extracting and quantifying nucleic acids simultaneously from these pathogen groups could enable cost-effective routine monitoring, offering invaluable insights into public health dynamics.

Understanding which components of the wastewater sample (e.g. solid, liquid, or whole influent) to analyze for optimal pathogen recovery is also essential for standardizing sample collection methods across laboratories. Previous research on partitioning experiments with endogenous and spiked-in viruses has shown that viruses partition to the solid phase when whole influent is separated by centrifugation.^7–10^ However, due to the low solid content of raw wastewater, most of the virus in a given volume may remain in the liquid phase, even after partitioning.^8,9^ As a result, studies have produced mixed findings regarding which sample component is best for monitoring viruses in wastewater. While some research has found similar or higher detection rates in solids,^11–14^ other studies have shown that liquid samples may have greater sensitivity.^9^ Similar studies comparing raw influent to primary settled solids have generally found higher virus concentration in the primary settled solids when compared on mass basis.^15–17^ To the best of our knowledge, no study has directly compared primary settled solids to centrifuged influent pellets, even though the two sample types are often used interchangeably for treatment facilities that cannot provide primary settled solids. Furthermore, no research has simultaneously compared sensitivity across all three sample types – matched primary settled solids, whole influent, and centrifuged influent pellets.

Here, we investigated trade-offs in pathogen detection and quantification of processing and analyzing settled solids, whole influent, and centrifuged influent. Our primary objectives were to identify reliable protocols for the simultaneous extraction of diverse microbial targets and to assess how sample type and normalization strategies influence detection outcomes. To this end, we evaluated multiple commercially available extraction kits using endogenous and externally added pathogens. We assessed reliability of detection across targets spanning DNA and RNA viruses (mpox, norovirus, SARS-CoV-2), bacteria (*Clostridium difficile, Campylobacter jejuni*), protozoan cysts (*Cryptosporidium* spp.), fungi (*Candida auris*), and clinically relevant antibiotic resistance genes (*CTX-M, mcr-1, NDM-1*, and *qnrS*). Additionally, we measured several endogenous and exogenous control viruses (bovine coronavirus, pepper mild mottle virus, and carjivirus, formerly known as crAssphage), and total bacteria (16S rRNA and rpoB genes). The chosen targets reflect a wide phenotypic diversity and aim to inform future wastewater monitoring efforts for public health. We then applied the best extraction methods to detect these targets in 24 solid and 32 liquid samples to compare detection rate and resulting concentrations. Finally, we investigated how different sample normalization strategies affected our results.

## 2. Materials & Methods

*Solid method development: sample collection, pre-analytical processing, and nucleic acid extraction* Grab samples of primary settled solids were collected from two wastewater treatment plants in California (Facility A and B) by the facility staff using sterile 50 mL falcon tubes.

Samples were transported on ice and held at 4□until analysis. Solids were dewatered by centrifugation at 20,000 xg for 30 minutes and 18 aliquots of approximately 0.25 g were transferred to sterile 2 mL microcentrifuge tubes. A cocktail of inactivated pathogens was prepared, which included mpox (Zeptometrix, NY), norovirus (Zeptometrix, NY), *Campylobacter jejuni* (Microbiologics, MN), *Clostridium difficile* (Microbiologics, MN), *Candida auris* (obtained from a collaborator), *Cryptosporidium parvum* (Microbiologics, MN), *Klebsiella pneumoniae* with plasmids containing sequences *CTX-M, NDM-1*, and *qnrS*, and plasmid containing *mcr-1* (made in-house; see SI). Bovine coronavirus (BCoV; Merck, NJ) served as an extraction control.

SARS-CoV-2 was not included due to the prevalence of COVID-19 in the communities at the time of the experiment (May - October 2023). Nine solids aliquots were spiked with the cocktail and vortexed gently. All samples were held at 4□ and nucleic acid extraction was performed within 24 hours. The moisture content of dewatered solids was determined by drying approximately 0.5 g of dewatered solids at 105□for 24 hours.

Nucleic acids from both the spiked and unspiked dewatered solids were extracted using three different kits, each in triplicate: the MagMAX Microbiome Ultra Nucleic Acid Isolation Kit (MagMAX; Thermo Fisher, MA), the AllPrep PowerViral DNA/RNA Kit (PowerViral; Qiagen, Germany), and the ZymoBIOMICS DNA/RNA Miniprep Kit (Zymo; Zymo Research, CA). All extractions were performed according to the manufacturers’ protocol with the following modifications: 1) The beads provided for bead beating in each kit were transferred to 2 mL tubes containing the aliquoted samples and the pathogen cocktail. 2) Each sample was bead-beaten for 20 minutes to enhance lysis, as recommended by the manufacturers. Extracted nucleic acids were stored at -80□until analysis. Each extraction included a reagent-only negative control.

### Influent method development: sample collection, pre-analytical processing, concentration, and nucleic acid extraction

Composite whole influent samples (1L) were collected from three wastewater treatment plants in California (Facility A, G, and I) by the facility staff using sterile 1L bottles. Samples were transported on ice and stored at 4 ºC until analysis. For each facility, 18 aliquots of approximately 35 mL were created, of which 9 were spiked with a cocktail of inactivated pathogens, consisting of mpox, *C. auris*, and *C. difficile*, with BCoV as an extraction control. The cocktail for Facility G additionally included *Cryptosporidium parvum*. However, after determining that our methods were capable of detecting endogenous *Cryptosporidium* spp. at measurable concentrations, this pathogen wasn’t included in the cocktails for Facility A and I. All other pathogens were detected in endogenous wastewater during solids methods testing, and assumed to be present. All aliquots were mixed on a rotating shaker for 3 hours at room temperature and then equilibrated at 4ºC. Extraction was performed within 24 hours.

All aliquots were concentrated and extracted using Promega Wizard Enviro TNA kit (Promega; Promega Corporation, WI) or Nanotrap Microbiome A and B particles (Ceres Nanosciences, VA), combined with AllPrep PowerViral DNA/RNA Kit (Nano-PowerViral) or ZymoBIOMICS DNA/RNA Miniprep Kit (Nano-Zymo) according to the manufacturer’s instructions. Extracted nucleic acids were stored at -80 ºC until analysis. Each extraction batch included a reagent-only negative control.

### Method application: sample collection & processing

1 L of whole influent and 50 mL of date-matched primary settled solids (when available) were collected in sterile bottles by facility staff from eleven facilities (nine wastewater treatment plants, one hospital, and one prison medical facility) on the West Coast (**Table S1**). Samples were either collected directly from nearby facilities or shipped overnight, kept on ice during transport, and processed within 48 hours of being received. Sample collection occurred at three time points: October-November of 2023, December 2023, and January 2024 (**Table S2**). For each time point, all samples were collected within three weeks of each other. Whole influent samples were divided into three 35 mL aliquots, spiked with BCoV, and processed using Nano-PowerViral. The remaining whole influent was aliquoted in 50 mL falcon tubes and centrifuged at 20,000 xg in 30 minute intervals and decanted to pellet the solids. Both centrifuged influent pellets and dewatered primary settled solids were aliquoted, spiked with BCoV, and processed using Zymo (additional details in SI).

### Nucleic acid quantification

Nucleic acids from all experiments were analyzed using the QIAcuity Four Digital PCR System (Qiagen, Germany). The QIAcuity OneStep Advanced Probe Kit (Qiagen, Germany) was used for RNA targets and the QIAcuity Probe PCR Kit (Qiagen, Germany) was used for DNA targets. For method application samples, we also utilized the QIAcuity EG PCR Kit (Qiagen, Germany) to measure 16S rRNA and rpoB. The assays used for each target are detailed in **Table S3**, and the cycling conditions are outlined in **Table S4**. Targets in high concentration (BCoV, carjivirus, PMMoV, norovirus, *CTX-M, qnrS*, 16S, rpoB) were detected using Qiacuity Nanoplates with 8.5k partitions, while targets in low concentration (*C. auris, C. difficile, C. jejuni, Cryptosporidium spp*., mpox, SARS-CoV-2, *NDM-1*, and *mcr-1*) were detected using plates with 26k partitions.

Each plate included positive controls (gBlocks containing the target sequences; IDT, IA) and negative controls (molecular-grade water). Concentration per reaction was converted to copies per gram of dry weight or copies per milliliter of wastewater using dimensional analysis (see SI). Where reported, inhibition tests were performed by comparing the resulting concentration of undiluted template to those diluted by 5-fold.

### Statistical analysis

Analysis was conducted using R (version 4.3.0). Difference in detection rate among primary settled solids, centrifuged influent pellets, and whole influent was assessed using the McNemar test for measurements that had all three sample types available. For subsequent analysis comparing two sample types, all available data that had a date-matched sample for the two sample types being analyzed was used. Nonparametric Kendall’s tau was used to assess correlation between measurements in different sample types as data were neither normally nor log-normally distributed based on Shapiro-Wilk tests. Correlation was assessed including the non-detects, which were set to 0. Linear regression of log-transformed concentrations was used to verify correlation between sample types and estimate the empirical relationship among measurements with detectable concentrations (i.e. non-detects excluded). Ratios of concentrations between two sample types were also calculated using measurements with detectable concentrations.

Our main analysis was conducted on a per mass basis, comparing target concentrations in copies per 1 g of solids to target concentrations in copies per 1 mL of liquid influent. We conducted a secondary analysis using concentration of the nucleic acid extracts (cp/uL), concentration on per volume basis (cp/mL), and concentrations normalized by our controls, PMMoV, carjivirus, 16S, and rpoB. Analysis for concentration on per-volume-basis used a subset of samples for which centrifuged influent volume was recorded. Dimensional analysis calculations are shown in the SI.

### QA/QC

All positive extraction and PCR controls were positive. For any negative extraction and PCR controls that were positive, we verified that the detected concentration in samples compared to the negative controls were at least one magnitude greater and therefore negligible. RNA targets were excluded as failed extractions if the samples were negative for BCoV or had fewer than 10^5^ cp/g dry weight for PMMoV, and DNA targets were excluded if the samples had fewer than 10^7^ cp/g dry weight for carjivirus. For wastewater influent samples, RNA targets were excluded as failed extractions if the samples were negative for BCoV or had fewer than 10^2^ cp/mL for PMMoV, and DNA targets were excluded if the samples had fewer than 10^4^ cp/mL for carjivirus.

## 3. Results

This study was conducted in two phases: first, we performed a methods comparison for each sample type (solids and whole influent); second, we applied the best performing methods to compare target concentrations in sample types across a large set of samples.

### 3.1 Method development

#### Solid samples

Detection rates for endogenous targets varied among the three extraction kits, and *C. difficile*, mpox, and *C. auris* were not detected in unspiked samples by any methods. PowerViral and Zymo showed similar rates of detection for most targets, while MagMAX performed particularly poorly for detecting *C. jejuni, NDM-1*, and *Cryptosporidium* spp. (**Figure 1**). In samples spiked with the pathogen cocktail, all targets were detected using all methods with a few exceptions for MagMAX (**Figure S1**).

**Figure 1.**
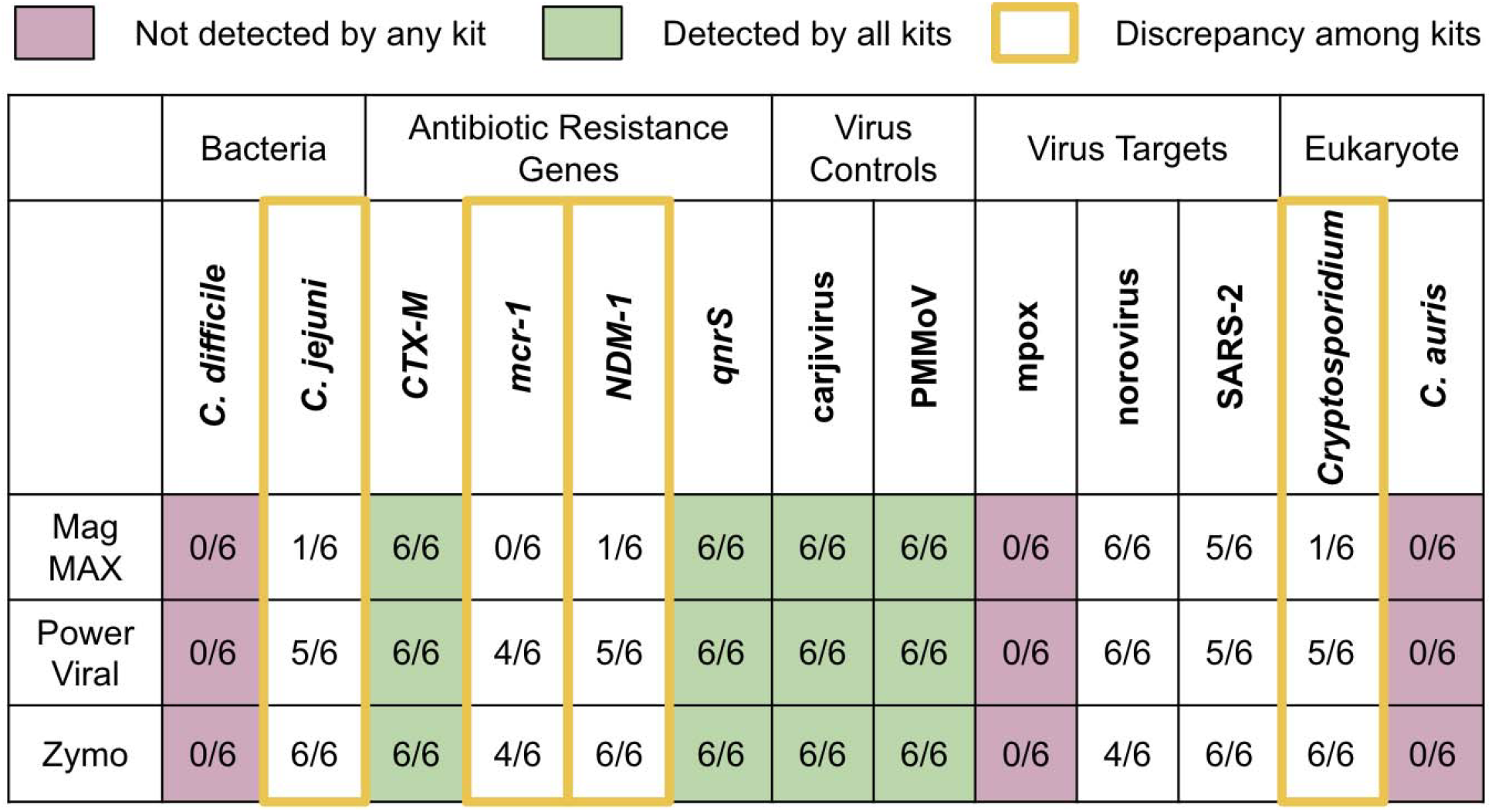
**Solid** methods comparison of **endogenous** targets. The detection rate is shown as the number of total replicates with measurable concentrations across two samples used for method comparison (n = 3 for each). Color indicates detection agreement level and yellow highlight shows targets with discrepancy between kits in greater than 50% of the replicates.

Next we compared concentrations of each target across methods measuring either endogenous and/or spiked-in targets (**Figure 2**). The concentrations of all targets from each extraction kit typically fell within one order of magnitude when detected. Although there were differences in endogenous target concentrations between samples from Facility A and B, the performance trends of the kits remained consistent between the two samples. With the exception of SARS-CoV-2, MagMAX yielded the highest concentration of viral targets. However, it produced the lowest concentration for some of the non-viral targets including *Cryptosporidium* spp., *C. difficile*, and *C. jejuni*, as well as bacteria with *mcr-1* and *NDM-1* genes. As MagMAX is widely used for viral targets in wastewater surveillance programs employing laboratory automation, we tested optimization methods such as heat pre-treatment in lysis buffer and bead beating using additional, larger beads to enhance lysis of larger targets. While these modifications had varying effects on different targets, they did not improve the detection of *Cryptosporidium* spp., one of the most challenging targets to detect in this study (**Figure S2A**). In contrast, PowerViral and Zymo performed well across all targets, but without clear trends. Zymo produced the highest concentration of *Cryptosporidium* spp. in both samples.

**Figure 2.**
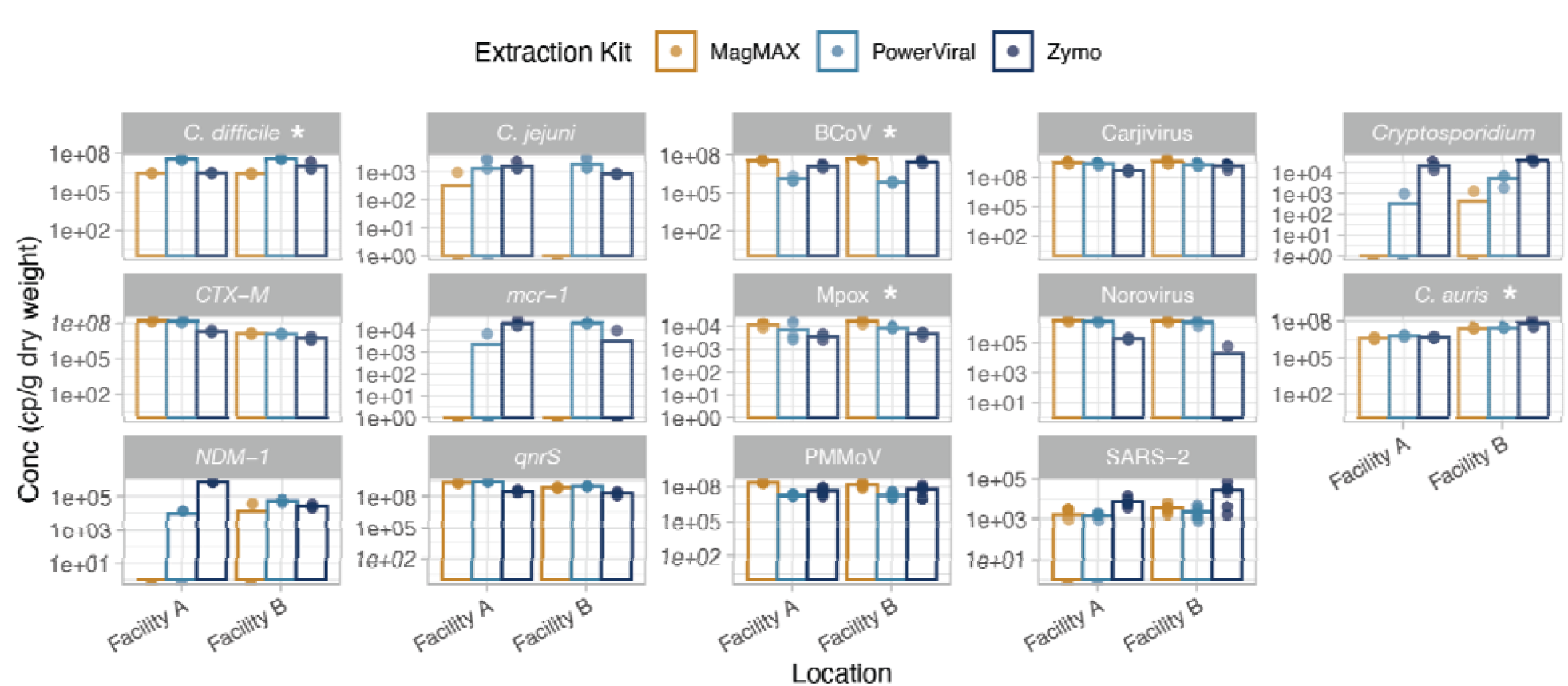
Comparison of three nucleic acid extraction kits on primary settled solids. Samples are from two wastewater treatment plants in California and results are shown for bacteria (1st and 2nd columns), virus (3rd and 4th columns), and eukaryotic targets (5th column). Each data point is a technical replicate (n = 3 for each facility) and the bar shows the average concentration. When undetected, the data point appears below the bar. Carjivirus, PMMoV, and SARS-2 were not spiked in any samples but measured across both spiked and unspiked samples (n = 6 for each facility). Note that y-axes differ by target and are log-scaled. *Spiked-in targets are indicated by an asterisk.

Nucleic acids extracted from each kit were tested for dPCR inhibition across three targets using a dilution method (**Figure S3**). Zymo consistently produced nucleic acids with low inhibition levels while MagMAX exhibited the highest inhibition. Modifications to MagMAX intended to mitigate inhibition, such as reducing the total input mass and increasing lysis buffer volume, did not yield improvements (**Figure S4B**). While nucleic acids from Facility A showed greater inhibition than those from Facility B across all kits, the trend among the three kits were similar. Prioritizing consistent detection of all targets, we selected Zymo to apply in a broad survey of samples.

#### Whole influent samples

Three concentration and extraction methods were compared using wastewater influent samples from three wastewater treatment plants: Promega Wizard Enviro TNA kit (Promega) and Nanotrap Microbiome A and B particles combined with either AllPrep PowerViral DNA/RNA Kit (Nano-PowerViral) or ZymoBIOMICS DNA/RNA Miniprep Kit (Nano-Zymo). The detection rates of endogenous targets varied among the three kits but overall, Nano-PowerViral showed superior detection compared to other kits (**Figure 3**). Endogenous *C. difficile*, mpox, and *C. auris* were not detected with any methods. Half of the aliquots were spiked with a cocktail containing these three targets and BCoV, and the endogenous concentrations of the other targets were also measured. In the spiked samples, most targets were detected using all three methods but Nano-Zymo had some difficulties with recovering mpox. (**Figure S5**).

**Figure 3.**
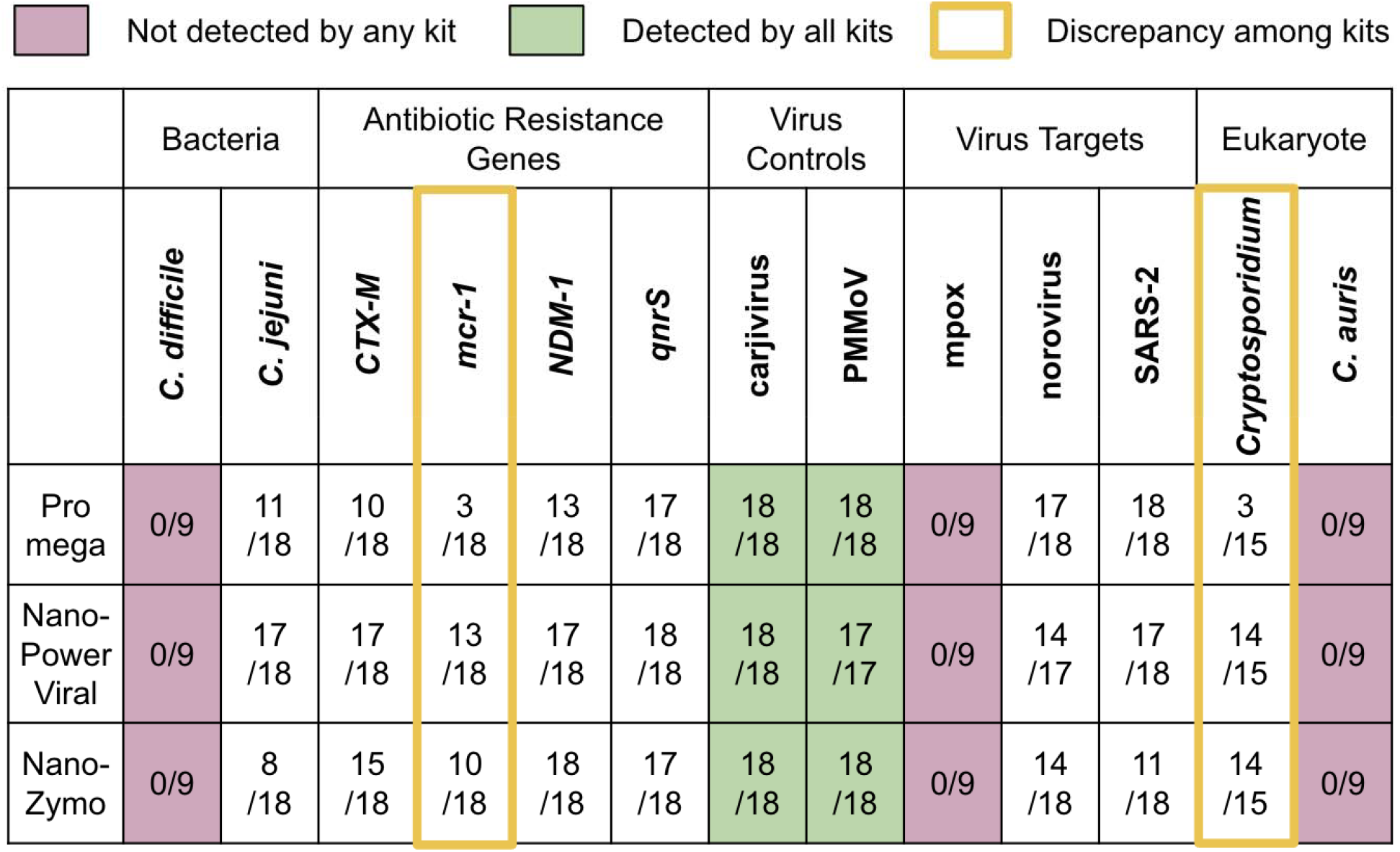
**Influent** methods comparison of **endogenous** targets. The detection rate is shown as the number of total replicates resulting with measurable concentration across samples used for method comparison. Color indicates detection agreement level and yellow highlight shows targets with discrepancy between kits in greater than 50% of the replicates.

For endogenous targets in wastewater, concentration comparison was conducted using data from both unspiked and spiked samples (n = 18). Data from the spiked sample was used for targets not expected to be prevalent in wastewater (n = 9), such as *C. difficile*, mpox, and *C. auris*. While both kits using Nanotrap Microbiome A and B particles (Nano-PowerViral, Nano-Zymo) exhibited comparable detection capabilities for most targets, Nano-PowerViral demonstrated enhanced performance by identifying higher concentrations of the targets and was therefore selected to apply in a broad survey of samples (**Figure 4**). Although inhibition testing was not conducted for whole influent, we expect the trend between the two extraction kits used for solids method development to behave similarly and for the inhibition rate to be lower than that of solids due to lower concentrations of inhibitory substances in whole influent.

**Figure 4.**
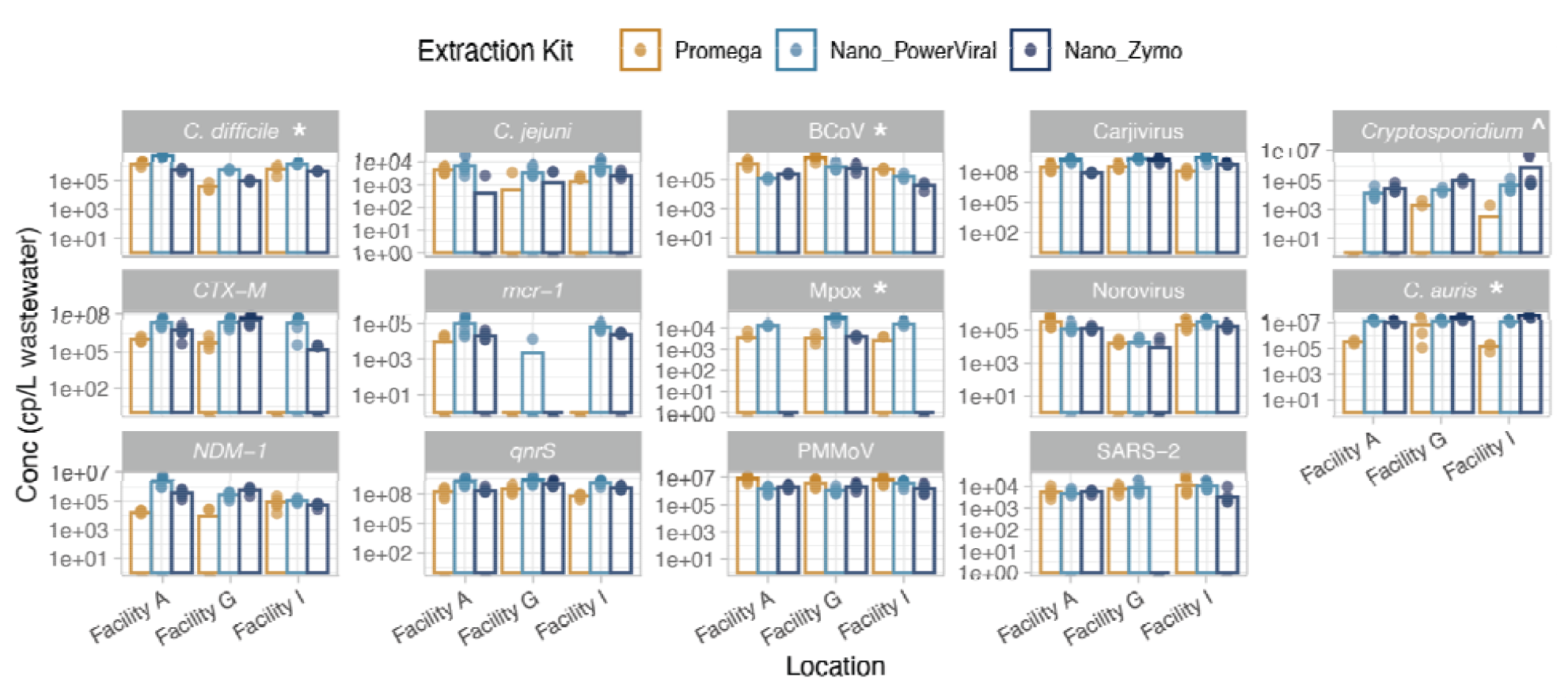
Comparison of three concentration/extraction methods on whole influent. Samples are from three wastewater treatment plants in California for bacteria (1st and 2nd columns), virus (3rd and 4th columns), and eukaryotic targets (5th column). Each data point is a technical replicate (n = 3 for spiked targets and n = 6 for unspiked targets per kit and facility) and the bar shows the average concentration. When undetected, the data point appears below the bar. Note that the y-axes differ by target and are log-scaled. *Spiked targets are indicated by an asterisk. ^^^Cryptosporidium was spiked for only Facility G (n = 5 per kit and facility).

### 3.2 Method application on wastewater from 11 sites

To understand the reliability and comparability of solids and whole influent methods, we quantified the same targets across whole influent and centrifuged influent pellet samples from 11 sites and date-matched primary settled solids samples from 8 of those sites at three different time points (**Table S2, Figure S6**). Target pathogens and controls were present in each sample type over a broad range of concentrations (**Figure S7**). Out of the 261 sample-by-target combinations with data from all three sample matrices (whole influent, centrifuged influent, and settled solids), 247 measurements were concordant among all matrices (i.e. all detects or all non-detects) and 14 measurements were discordant (i.e. only one or two sample types resulted in a detectable concentration of the target; **Figure 5A**). Targets with discordant measurements included *C. difficile, C. jejuni, mcr-1, Cryptosporidium* spp., *C. auris*, mpox, and SARS-CoV-2 (**Figure 5B**). Although centrifuged influent had a slightly higher detection rate, the difference in detection rates among the sample matrices was not significant when comparing two matrices at a time (Mcnemar test for solid vs. centrifuged influent: p = 0.29; centrifuged influent vs. influent: p = 0.29; influent vs. solid: p = 1).

**Figure 5.**
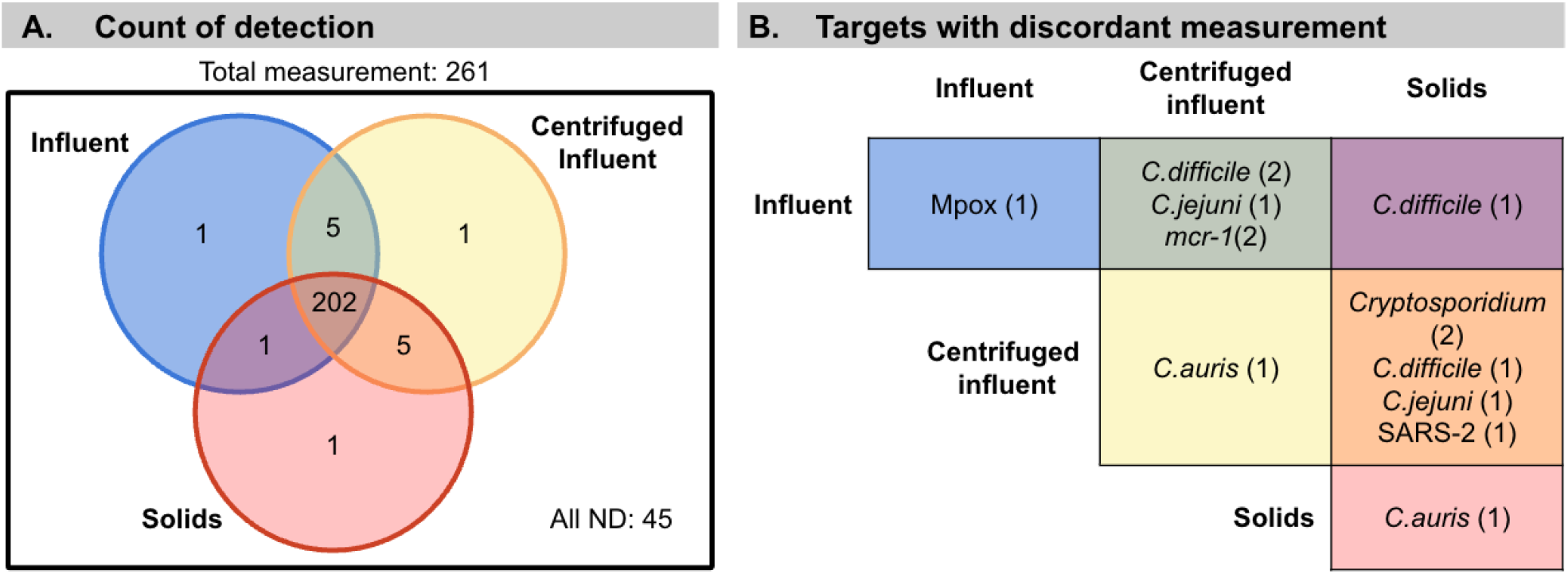
**A**) Count of sample-by-target measurements for 11 targets (excluding controls) across 24 samples that were detected in one, two, or all three sample types. “All ND” indicates combinations not detected by any method. Samples shown include only those from the eight sites that provided paired primary settled solids and whole influent. **B**) Table showing which matrices had a detectable measurement of the target indicated when at least one other sample matrix type was non-detect (discordant measurements). Rows and columns are matrices that the target was detected in and the color shows the corresponding area of the Venn diagram in A. The number of samples with each target detected is indicated in parentheses.

Quantitative measurements of sample-by-targets on per-mass-basis were positively and significantly correlated between sample matrices (**Figure 6A**; Kendall’s tau > 0.76, p < 0.001 for all matrix-by-matrix correlations). Linear regression performed on log_10_-transformed concentrations showed that for one log_10_ increase in any sample type, there was approximately one log_10_ increase in another (**Table S5**). At the individual target level, correlations were less consistent, potentially due to the small sample size and concentrations that varied over a narrow range. Kendall’s tau values for these individual comparisons ranged from no significant correlation to 0.67 between centrifuged influent and influent, 0.32 to 0.59 between primary settled solids and centrifuged influent, and no significant correlation to 0.68 between primary settled solids and influent (p < 0.05; **Table S6**). Centrifuged influent and primary settled solids had the highest number of targets with significant correlations, followed by centrifuged influent and influent (**Table S6**). On a per-mass-basis (g to mL), the median concentration ratios for all targets were as follows: centrifuged influent to influent was 3300 mL/g, primary solids to influent was 540 mL/g, and centrifuged influent to primary solids was 4 g/g (**Figure S8A, Table S7**).

**Figure 6.**
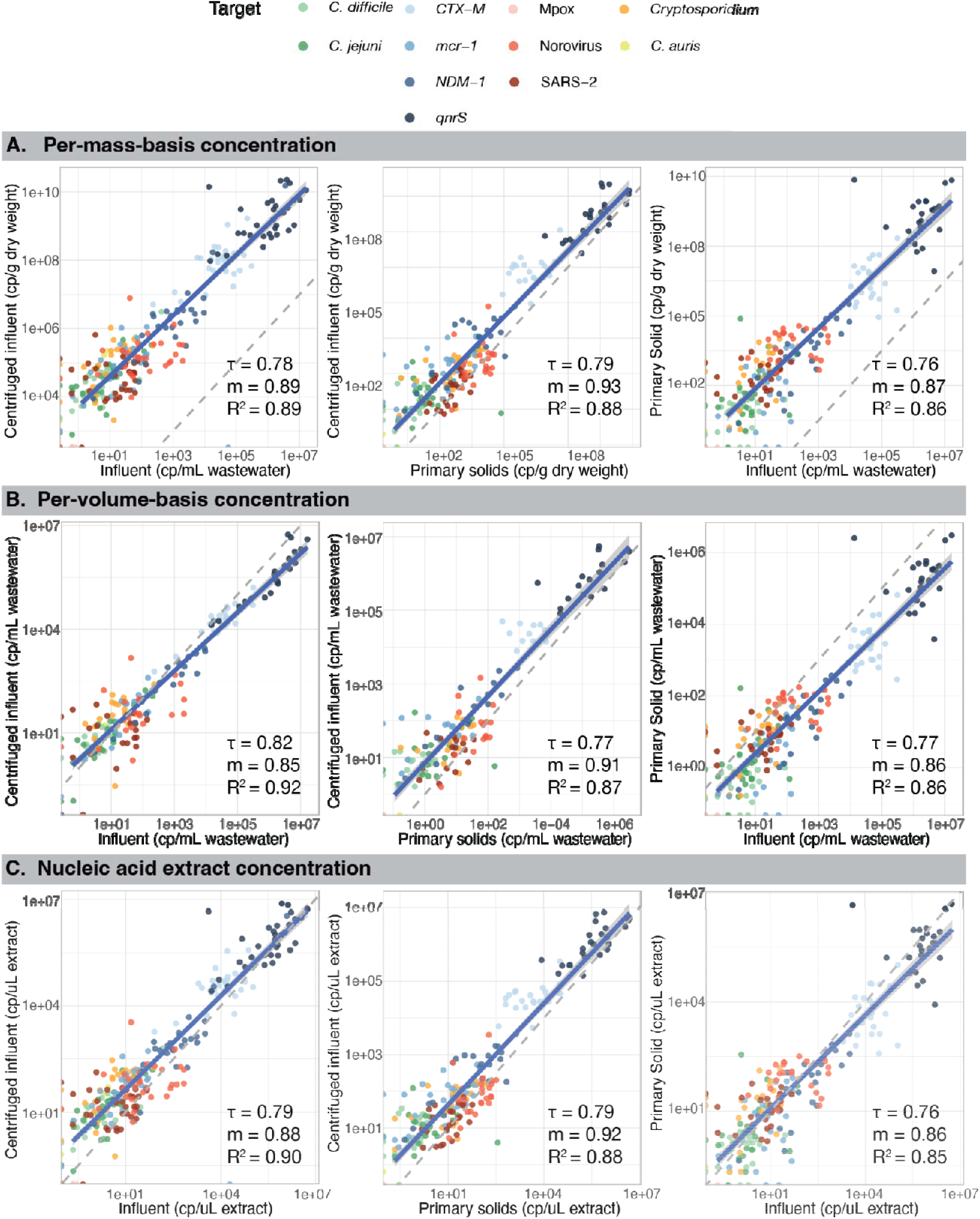
Comparison of wastewater concentrations by **A)** per-mass-basis, **B)** nucleic acid extract, and **C)** per-volume-basis of pathogen targets in paired samples processed by the three sample types. For comparisons with primary settled solids, samples are shown for only the eight wastewater treatment plants that provided paired primary settled solids. Points represent individual targets from one of the three time points at each location. Solid blue line shows linear regression with confidence interval for points that resulted in a measurable concentration, while the gray dashed line shows 1:1 relationship. Note log_10_-scale on both axes. Correlation coefficient Kendall’s tau and R^2^ of the linear regressions shown for each plot.

Most analysis in this study was conducted with raw concentration on a per-mass-basis (i.e. cp/mL wastewater and cp/g dry weight of solids). Secondary analyses included comparing concentrations using 1) per-volume-basis concentration, 2) in the extracted nucleic acids, and 3) normalizing by various controls. Analysis using per-volume-basis concentration resulted in similar correlations when all targets were included (**Figure 6B**; Kendall’s tau > 0.77, p < 0.001 for all matrix-by-matrix correlations). However the median ratios between sample types were 0.8:1 for centrifuged influent to influent, 5:1 for centrifuged influent to primary solids, and 1:6 for primary solids to influent (**Figure S8B, Table S7**). In nucleic acid concentrations, the correlations of the overall concentrations again remained the same (**Figure 6C;** Kendall’s tau > 0.76, p < 0.001 for all matrix-by-matrix correlations) and the median ratios between sample types were close to 4:1 for centrifuged influent to influent, 3.5:1 for centrifuged influent to primary solids, and 0.9:1 for primary solids to influent (**Figure S8C, Table S7**).

When data was normalized by PMMoV for RNA, carjivirus for DNA, and 16S for ARG targets, the correlations of overall concentrations were similar to the per mass correlation (**Figure S9A;** Kendall’s tau > 0.76, p < 0.001 for all matrix-by-matrix correlations). However, the median ratio was lower: 0.6:1 for centrifuged influent to influent, 1:1 for centrifuged influent to primary solids, and 0.4:1 for primary solids to influent (**Figure S10A**). When normalized with PMMoV for all targets, the overall correlation did not change significantly (**Figure S9B**; Kendall’s tau >0.7, p < 0.001 for all matrix-by-matrix correlations). However, because PMMoV concentrations were lower than the other controls (**Figure S7B**), the target concentrations were higher when normalized by PMMoV (**Figure S10B**). Concentrations normalized by 16S rRNA gene and rpoB gene (RNA polymerase single copy gene) correlated very well (**Figure S11;** Kendall’s tau > 0.91, p < 0.001 for all matrix-by-matrix correlation).

## 4 Discussion

For both solids and whole influent samples, each commercial extraction kit tested in this study demonstrated superior performance for specific targets, but no kit was the best across all targets. Our results indicate the optimal extraction method depends on the desired targets of a surveillance program. A tradeoff was observed between high recovery of viruses and organisms with more complex structures. For the solid samples, the ZymoBIOMICS DNA/RNA Miniprep Kit (Zymo) exhibited the most consistent performance and the lowest overall level of PCR inhibition. While it did not always yield the highest quantities of all targets compared to other kits, it successfully detected every target. In contrast, the MagMAX Microbiome Ultra Nucleic Acid Isolation kit (MagMAX) performed the worst overall, despite excelling with most viral targets.

Issues with inconsistent target detection were observed with this kit, potentially due to incomplete lysis or high levels of PCR inhibition. Additionally, manual isolation of nucleic acid conducted with a magnetic rack often resulted in poor separation of magnetic beads, which could have reduced recovery of nucleic acids.

Similarly, no single kit excelled for all targets in liquid samples. The Promega Wizard Enviro TNA kit (Promega), a direct extraction technique, performed well for viral targets but poorly for others. The lower recoveries were likely due to a combination of poor lysis of bacterial and eukaryotic targets by proteases and the fact that the intact organisms were pelleted by centrifugation during the solids removal step. CERES nanotrap A&B beads, which can potentially recover targets present in wastewater liquids and solids, coupled with either AllPrep PowerViral DNA/RNA (PowerViral) or Zymo, provided good performance across targets, with PowerViral yielding higher concentrations for some. MagMAX and Promega (commonly used for viral targets) showed poor performance for some bacterial and eukaryotic targets; others have also found that magnetic-bead based kits produce the lowest PCR diagnostic sensitivity.^18^ This highlights the need to optimize magnetic bead-based methods or shift to alternative methods to move towards an approach that detects a broader range of pathogens.

Our results demonstrate that a single workflow can be used to detect viral, bacterial, and eukaryotic targets. When the two selected extraction methods (Zymo for solids and Nano-PowerViral for liquids) were used with wastewater from 11 sites, all targets were detected except mpox, which was at low prevalence at the time of sample collection. Detection levels were similar for whole influent, centrifuged influent solids, and primary settled solids samples and the concentrations for all three sample types were strongly positively correlated. Thus, any of these sample types is likely viable for detecting a wide range of viruses, bacteria, protozoan cysts, fungi, and ARGs. On a per mass basis, centrifuged influent pellets were more concentrated than whole influent samples, with median ratios ranging from 1200 to 27000 mL/g for different targets and primary settled solids were more concentrated than whole influent samples, with median ratios ranging from 240 to 1600 mL/g. These are the first results published for *C. difficile, C. jejuni*, antibiotic resistance genes, *Cryptosporidium*, and *C. auris*.

The median ratios for viral targets SARS-CoV-2 and norovirus were lower than previously published values,^7,8,17^ potentially due to modifications aimed at improving the recovery of organisms with more complex structures, which may have reduced the efficiency for recovery of viruses.

When analyzed on a per-volume basis, influent samples exhibited higher concentrations than solid samples, due to the larger volume of wastewater associated with both centrifuged influent pellets and primary solids. Influent samples were processed in 35 mL aliquots, while 150 mL to 1 L of wastewater were associated with centrifuged influent pellets and 350 mL to 1.2 L with primary solids for each sample. The concentration of nucleic acids in the extract from each method provided insight that the sensitivity was comparable across sample types, with centrifuged influent showing slightly higher sensitivity. Therefore, processing centrifuged influent is a suitable alternative when sampling from primary clarifiers is not feasible or when sampling at manholes.

Centrifuged influent pellets generally resulted in higher concentration than primary settled solids. Centrifuged influent pellets effectively capture both pathogens naturally partitioned to the solid phase and those pelleted at high centrifugation speeds, regardless of their association with solids. In contrast, primary settled solids contain pathogens associated with solids that settle during primary sedimentation. As such, the median ratio of centrifuged influent to primary solids, with the exception of norovirus, was all greater than 1, showing effectiveness of capturing our targets in a pellet form (**Figure S8C**). None of the pathogen targets, including *Cryptosporidium* oocysts, are expected to settle during the settling times typical in mechanical treatment plants on their own. While oocysts attached to suspended particles have been shown to have a settling rate similar to other particles^19^ and have a higher settling rate with increasing particle size,^20^ the expected time to sediment completely still exceeds the typical settling time of most primary clarifiers. This is consistent with our finding that centrifuged influent pellets had the highest concentration of *Cryptosporidium* and whole influent the lowest, again demonstrating the benefit of high speed centrifugation to pellet targets.

Additionally, centrifuged influent addresses some of the limitations of primary settled solids, particularly the variable residence time of solids in primary clarifiers across facilities, which complicates linking a given sample to time of collection.^21^ Using 24-hour composite influent samples to obtain solids is often more intuitive and easier to interpret. However, centrifuging wastewater can be both time-consuming and labor intensive. Due to variability in total suspended solids concentrations in this study, different volumes of wastewater, ranging from 500 mL to 1 L, needed to be centrifuged to obtain a sufficient pellet. There is currently no standardized procedure for centrifuging wastewater to obtain solids for pathogen quantification. Previous studies have used a range of centrifugation speeds and durations, from 24,000 xg for 30 minutes^17^ to 1,840 xg for 30 minutes^13^ and 4,500 xg for 10 minutes.^11^ Some evidence suggests that higher centrifugation speeds and longer durations may increase viral RNA yield in the resulting pellet,^22^ highlighting the need for further optimization and standardization of centrifugation protocols.

We assessed normalization of targets using various controls. While PMMoV is commonly used for viral RNA targets and has potential utility in assessing fecal strength of the wastewater, its suitability for non-viral targets is questionable. Differences in capsid structure and genome type (RNA vs. DNA) can influence decisions for downstream methods (e.g. extraction and quantification). Additionally, studies have shown that when samples are centrifuged, PMMoV may partition differently from SARS-CoV-2,^11–13,22^ raising concerns about its use as an endogenous reference for normalizing targets, especially in solid samples. To normalize DNA targets, we used carjivirus, but further investigation is needed to confirm its appropriateness for non-viral DNA targets. Bacterial 16S rRNA gene was used to normalize ARG concentrations, as is standard practice for ARGs; however, because 16S copy number varies across organisms, we also used the single-copy gene rpoB, which can provide a more accurate ARG per cell equivalent estimate.^23^ The two normalized ARG concentrations showed excellent correlation, suggesting either can be used as an effective normalizer for ARGs.

Normalizing targets with different controls can introduce bias, particularly when the control concentrations differ significantly. Here, PMMoV had the lowest concentration in our samples compared to other controls, resulting in higher normalized concentrations for viral targets compared to those normalized by carjivirus or 16S, which was inconsistent with our actual measurements. A recent preprint by Wang et al. showed that relative recovery efficiency of PMMoV and other controls, such as carjivirus, can vary by method.^24^ Therefore, quantitative comparison across targets normalized by different controls may not be appropriate.

A limitation for our study is that the comparison of sample matrices is likely to be method-specific, as the effects of partitioning cannot be entirely separated from method effects. While we tested and identified the best method from a few commercially available kits for each matrix, it is possible that other methods that were not tested in this study might yield better results for specific targets or specific sample types. Additionally, our extractions were performed manually, but MagMAX and PowerViral can be automated on the KingFisher Flex System and Qiacube Connect, respectively. Results could vary if strength of magnets used for magnetic bead pulldown differ. Another limitation was use of spiked-in targets in evaluating the different extraction methods. Some studies have found that seeded targets can behave differently as they may not associate with solids as readily as targets that were excreted through feces.^11,22,25^ To alleviate this effect, we equilibrated our samples after spiking, which Roldan et al. found to be an effective way of ensuring that spiked viruses give a valid assessment of endogenous virus partitioning.^8^

Here, we developed two workflows that allowed us to simultaneously detect DNA and RNA of viral, bacterial, and eukaryotic targets from real world solid and liquid samples. From a public health perspective, there is a need to be able to conduct wastewater based surveillance for diverse phenotypes of organisms present in wastewater that have limited epidemiological surveillance, a high disease burden, and/or are vaccine preventable. Our results suggest that there is minimal practical difference in detection rate or detection limit among sample types for the targets examined in this study. Therefore, our results show that primary settled solids, whole influent, and centrifuged influent solids may all be used effectively to detect a broad range of targets, expanding utility and potential of wastewater surveillance.

## Data Availability

All data produced in the present study are available upon reasonable request to the authors

## 5 Acknowledgements

This work was supported by the U.S. Centers for Disease Control BAA (75D301-22-R-72097). We thank our collaborators Katie Scott and Matthew F. Blair at Virginia Tech for providing us with inactivated *Candida auris*. We are grateful to collaborating wastewater agencies and individuals at UNLV and California Department of Corrections and Rehabilitation for help with sample collection and their expertise on individual systems. Graphical abstract was generated with BioRender. AJP is a Chan Zuckerberg Biohub San Francisco Investigator.

## References

1. Tonkin, E. et al. Testing delay in an environment of low COVID-19 prevalence: A qualitative study of testing behaviour amongst symptomatic South Australians. SSM - Qual. Res. Health 2, 100099 (2022).

2. Wu, S. L. et al. Substantial underestimation of SARS-CoV-2 infection in the United States. Nat. Commun. 11, 4507 (2020).

3. Jajosky, R. A. & Groseclose, S. L. Evaluation of reporting timeliness of public health surveillance systems for infectious diseases. BMC Public Health 4, 29 (2004).

4. Zhang, S. et al. Molecular Methods for Pathogenic Bacteria Detection and Recent Advances in Wastewater Analysis. Water 13, 3551 (2021).

5. Zahedi, A., Monis, P., Deere, D. & Ryan, U. Wastewater-based epidemiology—surveillance and early detection of waterborne pathogens with a focus on SARS-CoV-2, Cryptosporidium and Giardia. Parasitol. Res. 120, 4167–4188 (2021).

6. Barber, C. et al. Community-Scale Wastewater Surveillance of Candida auris during an Ongoing Outbreak in Southern Nevada. Environ. Sci. Technol. 57, 1755–1763 (2023).

7. Roldan-Hernandez, L., Van Oost, C. & Boehm, A. B. Solid–liquid partitioning of dengue, West Nile, Zika, hepatitis A, influenza A, and SARS-CoV-2 viruses in wastewater from across the USA. Environ. Sci. Water Res. Technol. 10.1039.D4EW00225C (2024) doi:10.1039/D4EW00225C.

8. Roldan-Hernandez, L. & Boehm, A. B. Adsorption of Respiratory Syncytial Virus, Rhinovirus, SARS-CoV-2, and F+ Bacteriophage MS2 RNA onto Wastewater Solids from Raw Wastewater. Environ. Sci. Technol. 57, 13346–13355 (2023).

9. Wu, J. et al. Multiplexed Detection, Partitioning, and Persistence of Wild-Type and Vaccine Strains of Measles, Mumps, and Rubella Viruses in Wastewater. Environ. Sci. Technol. 58, 21930–21941 (2024).

10. Mercier, E. et al. Municipal and neighbourhood level wastewater surveillance and subtyping of an influenza virus outbreak. Sci. Rep. 12, 15777 (2022).

11. Hasing, M. et al. Comparison of Detecting and Quantitating SARS-CoV-2 in Wastewater Using Moderate-Speed Centrifuged Solids versus an Ultrafiltration Method. Water 13, 2166 (2021).

12. Kitakawa, K., Kitamura, K. & Yoshida, H. Monitoring Enteroviruses and SARS-CoV-2 in Wastewater Using the Polio Environmental Surveillance System in Japan. Appl. Environ. Microbiol. 89, e01853–22 (2023).

13. Kitamura, K., Sadamasu, K., Muramatsu, M. & Yoshida, H. Efficient detection of SARS-CoV-2 RNA in the solid fraction of wastewater. Sci. Total Environ. 763, 144587 (2021).

14. Tomasino, M. P. et al. SARS-CoV-2 RNA detected in urban wastewater from Porto, Portugal: Method optimization and continuous 25-week monitoring. Sci. Total Environ. 792, 148467 (2021).

15. Wolfe, M. K. et al. Wastewater-Based Detection of Two Influenza Outbreaks. Environ. Sci. Technol. Lett. 9, 687–692 (2022).

16. Kim, S. et al. SARS-CoV-2 RNA is enriched by orders of magnitude in primary settled solids relative to liquid wastewater at publicly owned treatment works. Environ. Sci. Water Res. Technol. 8, 757–770 (2022).

17. Boehm, A. B. et al. A retrospective longitudinal study of adenovirus group F, norovirus GI and GII, rotavirus, and enterovirus nucleic acids in wastewater solids at two wastewater treatment plants: solid-liquid partitioning and relation to clinical testing data. mSphere 9, e00736–23 (2024).

18. Paulos, S. et al. Evaluation of five commercial methods for the extraction and purification of DNA from human faecal samples for downstream molecular detection of the enteric protozoan parasites Cryptosporidium spp., Giardia duodenalis, and Entamoeba spp. J. Microbiol. Methods 127, 68–73 (2016).

19. Searcy, K. E., Packman, A. I., Atwill, E. R. & Harter, T. Association of Cryptosporidium parvum with Suspended Particles: Impact on Oocyst Sedimentation. Appl. Environ. Microbiol. 71, 1072–1078 (2005).

20. Medema, G. J., Schets, F. M., Teunis, P. F. M. & Havelaar, A. H. Sedimentation of Free and Attached Cryptosporidium Oocysts and Giardia Cysts in Water. Appl. Environ. Microbiol. 64, 4460–4466 (1998).

21. Wigginton, K. R. NSF Research Coordination Network Webinar Series - Lightening talks. (2024).

22. Breadner, P. R. et al. A comparative analysis of the partitioning behaviour of SARS-CoV-2 RNA in liquid and solid fractions of wastewater. Sci. Total Environ. 895, 165095 (2023).

23. Dahllöf, I., Baillie, H. & Kjelleberg, S. rpoB-Based Microbial Community Analysis Avoids Limitations Inherent in 16S rRNA Gene Intraspecies Heterogeneity. Appl. Environ. Microbiol. 66, 3376–3380 (2000).

24. Wang, A. L. et al. Benchmarking concentration and direct extraction methods for wastewater-based surveillance of eight human respiratory viruses: implications for rapid application to novel pathogens. Preprint at 10.1101/2024.11.27.625007 (2024).

25. Chik, A. H. S. et al. Comparison of approaches to quantify SARS-CoV-2 in wastewater using RT-qPCR: Results and implications from a collaborative inter-laboratory study in Canada. J. Environ. Sci. 107, 218–229 (2021).

