## Supplemental Information for "Solid evidence and liquid gold: trade-offs of processing settled solids, whole influent, or centrifuged influent for co-detecting viral, bacterial, and eukaryotic pathogens in wastewater"

*co-first authors

Affiliation

### Supporting Information

**Information on extraction kits used in this study**

MagMAX Microbiome Ultra Nucleic Acid Isolation Kit (MagMAX; Thermo Fisher, MA) employs a magnetic bead-based extraction method, whereas AllPrep PowerViral DNA/RNA Kit (PowerViral; Qiagen, Germany), and the ZymoBIOMICS DNA/RNA Miniprep Kit (Zymo; Zymo Research, CA) utilize silica column-based extraction. Promega Wizard Enviro TNA kit (Promega; Promega Corporation, WI) is a simultaneous concentration and extraction kit that begins with lysis and direct capture of nucleic acids on silica resin, followed by the addition of wash buffers to remove contaminants. Nanotrap Microbiome A and B particles (Ceres Nanosciences, VA) are hydrogel particles with chemical affinity baits that allow selective capture and enrichment of various targets.

**Additional details on method application**

Upon arrival, the whole influent samples were divided into three 35 mL aliquots, spiked with BCoV, and processed using the best performing liquid method determined in the method development experiment above. Briefly, the samples were equilibrated for 3-24 hours on a rotating shaker. Ceres Nanotrap Microbiome A & B beads together with Allprep PowerViral DNA/RNA Kit were used to concentrate and extract nucleic acids. The remaining whole influent was aliquoted in 50 mL falcon tubes and centrifuged at 20,000 xg in 30 minute intervals and decanted to pellet the solids. The volume of influent centrifuged varied depending on the solid content but typically ranged from 500 mL to 1 L to obtain sufficient mass for three replicates and a moisture content measurement. If there were not enough solids for triplicates, either duplicate or a single sample was prepped with a minimal amount for moisture content measurement. During the second and third collection periods, we measured the total weight of the centrifuged influent to determine the volume concentrated to obtain the pellet.

The date-matched primary settled solids were centrifuged at 20,000 xg in 50 mL falcon tubes for 30 minutes and decanted. Both centrifuged influent pellets and dewatered primary settled solids were aliquoted, spiked with BCoV, equilibrated for 1-24 hours at 4 ºC, and processed using the best performing solid method selected from the experiment described above. Briefly, the samples were bead-beaten for 20 minutes at maximum speed on a vortex using the beads in ZymoBIOMICS DNA/RNA Miniprep Kit and processed with modifications mentioned above. DNA and RNA were extracted separately to maximize yield. Resulting nucleic acid was stored at -80 ºC until analysis. Each extraction included a negative extraction control to detect potential contamination.

Data were analyzed with the QIAcuity Software Suite (version 2.5.0.1; Qiagen, Germany) and manually thresholded. A measurement was considered positive if at least three positive partitions were detected. If more than 80% of valid wells were positive, the sample was deemed oversaturated and was rerun with higher dilution. Replicates were averaged to calculate a mean concentration for the target being measured. A measurement was considered positive if at least one of the replicates had a measurable concentration.

**Methods for antibiotic resistance gene spike-ins**

Two positive controls were used for the antibiotic resistance gene (ARG) spike-in experiments. A *Klebsiella pneumoniae* isolate containing *CTX-M*, *NDM-1*, and *qnrS* genes from the CDC Antimicrobial Resistance Isolate Bank Enterobacterales Carbapenemase Diversity (CRE) panel (AR Bank Isolate #0145, SRA #SAMN04014986) was used as one ARG standard. Briefly, 20 mL of LB broth was inoculated with 20 μL of the isolate stock and incubated at 37℃ for 18 hours. Aliquots of 2 mL of broth were centrifuged at 5,000 xg followed by removal of the supernatant. The resulting pellet was heat inactivated at 80 ºC for 20 minutes. For the *mcr-1* positive control, an *mcr-1* DNA insert sequence was synthesized by IDT in a pUCIDT-AMP GoldenGate+ vector.

**MagMAX Microbiome Ultra Nucleic Acid Isolation Kit modifications**

Two sets of modifications were made: the first set was aiming to improve lysis as MM seem to do poorly with hard-to-lyse targets like *Cryptosporidium*. The second set was aimed to alleviate inhibition as MM had higher inhibition rates than the other two kits tested. For improving lysis, we tested two modifications in triplicate: 1) adding an additional heat lysis step before bead beating at 65 ºC for 10 minutes, and 2) adding additional stainless steel 5/32” grinding balls (OPS Diagnostics, NJ) to the bead beating step with the original beads. For this set, inactivated BCoV, Mpox, *C. difficile*, and *C. auris* was spiked in as a pathogen cocktail in equal amounts to all aliquots of primary settled solids from Plant B. The modifications did not result in improvement in detection of the targets. None, including the baseline extraction method (which followed the soil protocol from the manufacturer), were able to detect *Cryptosporidium* in the sample. While there was marginal improvement in recovery of *C. difficile*, there was a drastic decrease in recovery of *C. jejuni*, as well as minor decreases in viral targets as well (**Figure S1**). These modifications did not result in less inhibition as all measurements were either too inhibited and detected in only the 5-fold dilution but not in undiluted or was not detected in either, resulting in inconclusive results for measuring inhibition (**Figure S2**).

To alleviate inhibition, we tested two main modifications: 1) adding extra lysis buffer and 2) decreasing sample input. The second modification was done by using the “fecal protocol” instead of the “soil protocol,” where the main difference is decrease in sample input (100 mg instead of 250 mg) and omission of proteinase K. In order to know how each of these affected our results, we also tested the soil protocol without proteinase K, which resulted in testing three modifications. For this set, only inactivated BCoV was spiked in equal amounts to all aliquots from the same settled solids sample from Plant B. This set of modifications did improve detection of Cryptosporidium somewhat but detection was still inconsistent with only some of the replicates resulting in a detect. Additionally, it decreased recovery of C. jejuni in terms of the resulting concentration and also the consistency of replicate performance (**Figure S3**). While these modifications were made to alleviate inhibition, when testing for inhibition, the results showed that they were not enough to make a significant difference. The measurements were still either too inhibited and detected in only the 5-fold dilution but not in undiluted or was not detected in either, resulting in inconclusive results for measuring inhibition (**Figure S4**).

**Dimensional analysis**

In order to convert from X copies/μL obtained with dPCR to Y copies/g dry weight for any solid samples the following equation was used:

$$X \frac{copies}{\mu L rxn} \times\frac{Volume of rxn (\mu L)}{Volume of template in rxn (\mu L)}\times dilution factor$$

$\times\frac{Volume of eluent from extract (\mu L)}{Wet mass of solids in extract (g)}\times\% solids of sample = Y \frac{copies}{g dry weight}$

To convert Y copies/g dry weight to Z copies/mL wastewater for primary settled solid samples, the following equation was used where TSS is the total suspended solids (mg/L) provided by the wastewater facility for the dates sampled:

$$Y \frac{copies}{g dry weight} \times TSS = Z\frac{copies}{mL wastewater}$$

In order to convert from X copies/μL obtained with dPCR to Y copies/mL of wastewater for liquid samples, the following equation was used for influent:

$$X \frac{copies}{\mu L rxn} \times\frac{Volume of rxn (\mu L)}{Volume of template in rxn (\mu L)}\times dilution factor$$

$$\times\frac{Volume of eluent from extract (\mu L)}{Volume of wastewater extracted (mL)}= Y \frac{copies}{mL wastewater}$$

In order to convert from X copies/μL obtained with dPCR to Y copies/mL of wastewater for centrifuged influent, the following equation was used:

$$X \frac{copies}{\mu L rxn} \times\frac{Volume of rxn (\mu L)}{Volume of template in rxn (\mu L)}\times dilution factor$$

$$\times\frac{Volume of eluent from extract (\mu L)}{Wet mass of solids in extract (g)}\times\frac{Mass of pellet (g)}{Volume of wastewater centrifuged (mL)}= Y \frac{copies}{mL wastewater}$$

**Table S1. Facilities involved in this study.** Location, population served, and pretreatment (if any) before raw influent is collected is noted, as well as whether it was possible to collect primary settled solids from each of the plant (Y: yes. N: no).

| **Facility** | **State** | **Population** | **Notes/Pretreatment** | **Solids** |
| --- | --- | --- | --- | --- |
| Publicly owned treatment works | | | | |
| A | California | 740,000 |  | Y |
| B | California | 480,000 |  | Y |
| C | California | 280,000 | Use of ferric chloride; waste stream that contains treated sludge returns to downstream of influent but upstream of primary settled solids if solids removal not operating efficiently | Y |
| D | California | 510,000 |  | Y |
| E | California | 4,000,000 |  | Y |
| F | California | 775,000 | Physical screening & chlorination for odor control | Y |
| G | California | 18,000 | Physical screening | Y |
| H | Washington | 735,800 | No pretreatment before sampling | N |
| I | California | 70,000 |  | Y |
| Smaller facilities | | | | |
| J | California | 2,172 | Prison medical facility | N |
| K | Nevada | N/A | Hospital | N |

**Table S2. Sampling detail on raw influent and primary settled solids when available.** Dates of sample collection for all three time points are listed. For NVH, the December sample was not able to be collected due to logistical challenges.

| **Facility** | **Influent** | **Solids** | **Sample dates** |
| --- | --- | --- | --- |
| A: | 24 hour composite |  | 10/25/2023, 12/04/2023,  01/24/2024 |
| B | 24 hour composite |  | 11/13/2023,  12/14/2023,  01/25/2024 |
| C | 24 hour composite |  | 10/25/2023,  12/13/2023  01/24/2024 |
| D | 24 hour composite collected from 0005 to 2359 | Grab sample collected between 0600 and 0700 | 11/15/2023,  12/07/2023,  01/24/2024 |
| E | 24 hour composite collected from 0100 to 2300 manually | Composited manually from 0000 to 2300 | 10/30/2023, 12/03/2023,  01/21/2024 |
| F | 24 hour composite collected from midnight to next midnight every 30 minutes |  | 10/24/2023,  12/06/2023,  01/23/2024 |
| G | 24 hour composite |  | 10/24/2023,  12/05/2023,  01/23/2024 |
| H | 24 hour composite | Not available | 10/26/2023,  12/06/2023,  01/24/2024 |
| I | 24 hour composite collected |  | 10/24/2023, 12/05/2023,  01/23/2024 |
| J | 24 hour composite collected from 0800 to 0800 as 100 mL every 15 minutes | Not available | 10/24/2023, 12/05/2023,  01/24/2024 |
| K | 24 hour composite | Not available | 10/31/2023,  01/24/2024 |

**Table S3. Assays used in this study to detect target organisms and antibiotic resistance genes (ARGs).** *Cryptosporidium assay was used as a multiplex assay with TAMRA and a singleplex assay with FAM.

| **Target Organism** | **Primer/Probe** | **Sequence (5’-3’)** | **Reference** |
| --- | --- | --- | --- |
| Bovine coronavirus (BCoV) | F | CTGGAAGTTGGTGGAGTT | ^26^ |
|  | R | ATTATCGGCCTAACATACATC |  |
|  | P | CCTTCATATCTATACACATCAAGTTGTT-TAMRA |  |
| Carjivirus (formerly known as crAssphage) | F | CAGAAGTACAAACTCCTAAAAAACGTAGAG | ^27^ |
|  | R | GATGACCAATAAACAAGCCATTAGC |  |
|  | P | AATAACGATTTACGTGATGTAAC-TexasRed |  |
| Pepper mild mottle virus (PMMoV) | F | GAGTGGTTTGACCTTAACGTTTGA | ^28^ |
|  | R | TTGTCGGTTGCAATGCAAGT |  |
|  | P | MGB-CCTACCGAAGCAAATG-FAM |  |
| *Campylobacter jejuni* | F | TCCAAAATCCTCACTTGCCATT | ^29^ |
|  | R | TGCACCAGTGACTATGAATAACGA |  |
|  | P | TGCAACCTCACTAGCAAAATCCACAGCT-Cy5 |  |
| *Candida auris* | F | CAGACGTGAATCATCGAATCT | ^30^ |
|  | R | TTTCGTGCAAGCTGTAATTT |  |
|  | P | AATCTTCGCGGTGGCGTTGCATTCA-TexasRed |  |
| *Clostridium difficile* | F | GGTATTACCTAATGCTCCAAATAG | ^31^ |
|  | R | TTTGTGCCATCATTTTCTAAGC |  |
|  | P | ACCTGGTGTCCATCCTGTTTCCCA-HEX |  |
| *Cryptosporidium spp.* | F | ATGACGGGTAACGGGGAAT | ^32^ |
|  | R | CCAATTACAAAACCAAAAAGTCC |  |
|  | P | CGCGCCTGCTGCCTTCCTTAGATG-TAMRA/FAM* |  |
| hMPXV | F | TCAACTGAAAAGGCCATCTATGA | ^33^ |
|  | R | GAGTATAGAGCACTATTTCTAAATCCCA |  |
|  | P | CCATGCAATATACGTACAAGATAGTAGCCAAC-FAM |  |
| Norovirus GII | F | ATGTTCAGRTGGATGAGRTTCTCWGA | ^34^ |
|  | R | TCGACGCCATCTTCATTCACA |  |
|  | P | AGCACGTGGGAGGGCGATCG-FAM |  |
| ARG: *CTX-M* | F | CCGTCACGCTGTTRTTAGGA | ^35^ |
|  | R | AATGCCACMCCCAGYCKKCC |  |
|  | P | CAGCAAAAACTTGCCGRATT-FAM |  |
| ARG: *mcr-1* | F | GATCGCTGTCGTGCTCTTTG | ^35^ |
|  | R | ACCGCGCCCATGATTAATAG |  |
|  | P | CGATGCTACTGATCACCACG-TexasRed |  |
| ARG: *NDM-1* | F | ATATCACCGTTGGGATCGAC | ^36^ |
|  | R | TAGTGCTCAGTGTCGGCATC |  |
|  | P | AAGGACAGCAAGGCCAAGTCG-HEX |  |
| ARG: *qnrS* | F | TTGCTCAGCMTTTATTWCWGGATGT | ^35^ |
|  | R | CAGCGATTTTCAWACARCTCACA |  |
|  | P | TATGCCAATATGGAGMGGGT-TAMRA |  |
| 16S rRNA | F | CGGTGAATACGTTCYCGG | ^37^ |
|  | R | GGWTACCTTGTTACGACTT |  |
| rpoB | F | CGAACATCGGTCTGATCAACTC | ^38^ |
|  | R | CGCTGCATGTTCGAACCCAT |  |

**Table S4. PCR cycling conditions used to quantify nucleic acids.**

| **Cycle step** | **Temp** | **Time** | **Cycle No.** |
| --- | --- | --- | --- |
| RNA: QIAcuity OneStep Advanced Probe Kit | | | |
| Reverse transcription | 50℃ | 40 min | 1 |
| RT enzyme inactivation | 95℃ | 2 min | 1 |
| Denaturation | 95℃ | 5 sec | 45 |
| Combined annealing/extension | 60℃ | 30 sec |  |
| DNA: QIAcuity Probe PCR Kit | | | |
| Heat activation | 95℃ | 2 min | 1 |
| Denaturation | 95℃ | 15 sec | 45 |
| Combined annealing/extension | 60℃ | 30 sec |  |
| EvaGreen: QIAcuity EG PCR Kit | | | |
| Heat activation | 95℃ | 2 min | 1 |
| Denaturation | 95℃ | 15s | 40 |
| Annealing | 55℃ | 15s |  |
| Extension | 72℃ | 15s |  |
| Cooling down | 40℃ | 5 min | 1 |

**Table S5**. **Empirical relationship between concentrations measured in influent, centrifuged influent, and primary settled solids.** Y = mx + b where x, y = log_10_ transformed concentration of targets, m = slope, and b = intercept.

|  | **m** | **b** | **Adjusted R^2^** | **p-value** |
| --- | --- | --- | --- | --- |
| **Centrifuged influent vs influent** | 0.89 | 3.72 | 0.89 | < 0.001 |
| **Centrifuged influent vs primary solids** | 0.93 | 1.03 | 0.88 | < 0.001 |
| **Primary solids vs influent** | 0.87 | 3.04 | 0.86 | < 0.001 |

**Table S6**. **Correlation coefficients Kendall’s tau and p-values for correlation between two sample types for targets detected in at least 3 samples of each matrix when looking at concentrations per mass basis** (p < 0.05; *C. auris* and mpox excluded due to insufficient detections). - shows correlations that were not statistically significant.

| **Targets** | **Centrifuged influent  vs influent** | | **Centrifuged influent  vs primary solids** | | **Primary solids  vs influent** | |
| --- | --- | --- | --- | --- | --- | --- |
|  | Kendall’s tau | p-value | Kendall’s tau | p-value | Kendall’s tau | p-value |
| *C. difficile* | 0.57 | < 0.001 | 0.32 | 0.031 | - | 0.143 |
| *C. jejuni* | 0.62 | < 0.001 | 0.33 | 0.023 | 0.34 | 0.020 |
| *CTX-M* | 0.29 | 0.021 | 0.36 | 0.013 | - | 0.47 |
| *mcr-1* | 0.47 | < 0.001 | 0.33 | 0.024 | - | 0.087 |
| *NDM-1* | 0.67 | < 0.001 | 0.48 | 0.001 | 0.68 | < 0.001 |
| *qnrS* | 0.42 | < 0.001 | 0.59 | < 0.001 | - | 0.14 |
| *Crypto sporidium* | 0.33 | 0.011 | 0.50 | < 0.001 | 0.32 | 0.030 |
| Norovirus | 0.25 | 0.046 | 0.32 | 0.030 | - | 0.64 |
| SARS-2 | - | 0.45 | 0.36 | 0.013 | - | 0.14 |

**Table S7**. **Concentration ratio for matched samples analyzed.**

|  | **Centrifuged influent : Influent** | **Centrifuged influent : Solid** | **Solid : Influent** |
| --- | --- | --- | --- |
| Per-mass-basis | 3300 mL/g | 4.1 g/g | 540 mL/g |
| Per-volume-basis | 0.83 | 4.3 | 0.16 |
| Extract concentration | 3.9 | 3.5 | 0.87 |
| Normalized by controls | 0.60 | 1.3 | 0.37 |
| Normalized by PMMoV | 0.19 | 1.4 | 0.10 |

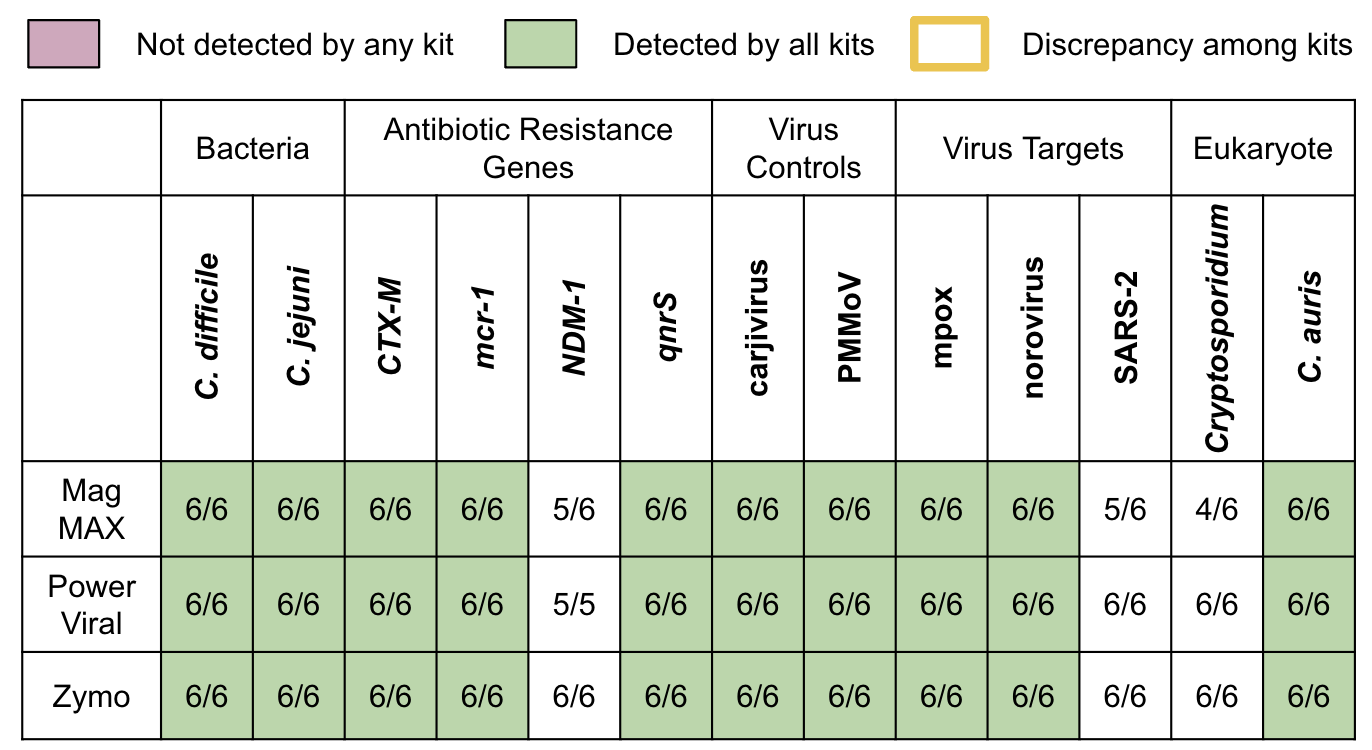

**Figure S1**. **Solid** methods comparison of **spiked-in** targets. The detection rate is shown as the number of total replicates with measurable concentrations across two samples used for method comparison (n = 3 for each). Color indicates detection agreement level.

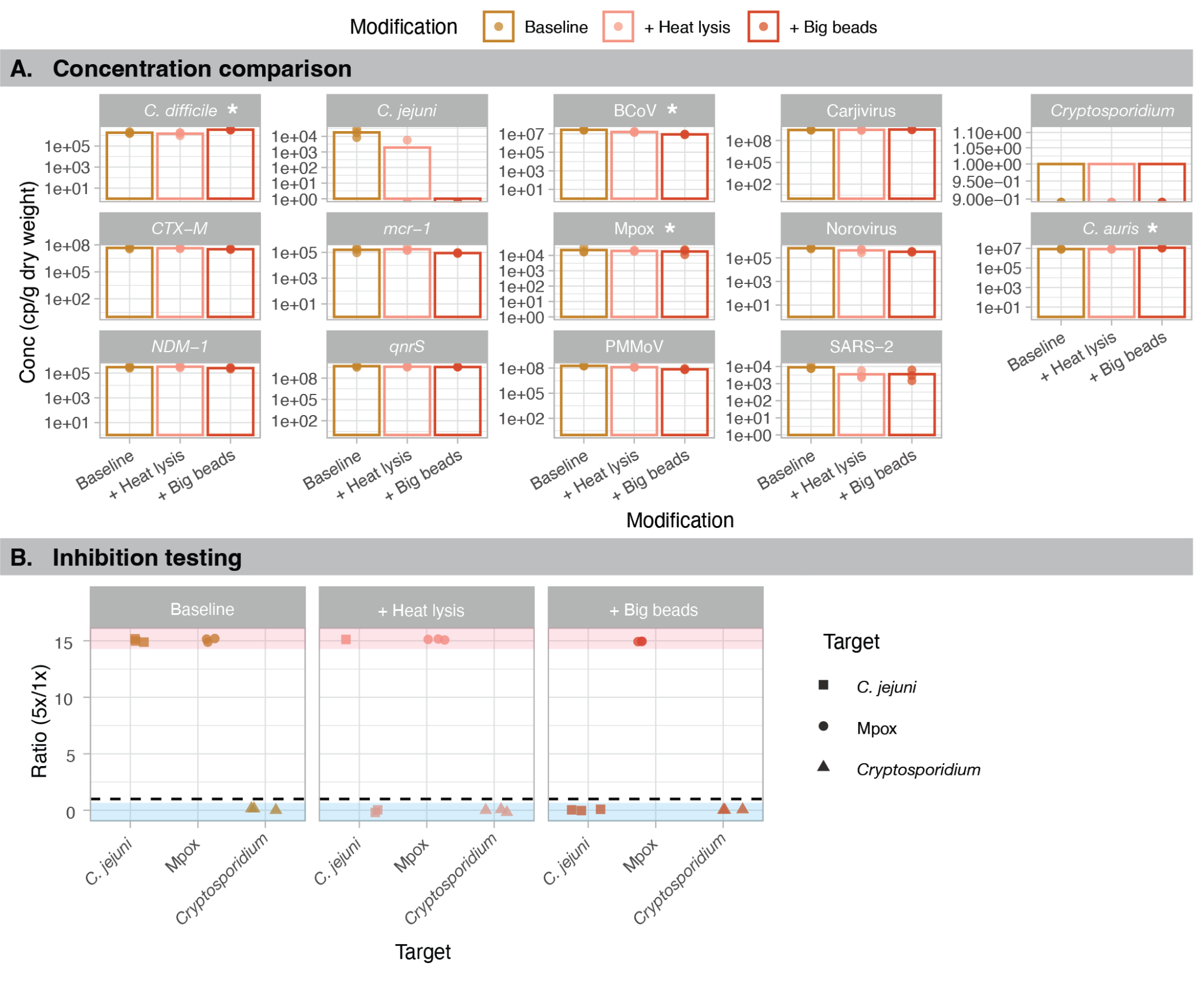

**Figure S2**. **A)** Concentration comparison of MagMAX Microbiome Ultra Nucleic Acid Isolation Kit (Baseline) with two modifications geared to enhance lysis with addition of heat lysis or bigger beads. Each data point is a technical replicate (n = 3) and the bar shows the average concentration. When undetected, the data point shows up under the bar graph. Note that y-axes differ by target and are log-scaled. *Spiked-in targets are indicated by an asterisk. **B)** dPCR inhibition for each modification of MagMAX geared for better lysis. Samples are considered inhibited if the ratio is greater than 1 (dashed line). Points located at the top of the plot in the red-shaded region (infinity) indicate that the sample was inhibited and the target was detected only in the 5-fold dilution but not in the undiluted template. Conversely, points at the bottom of the plot in the blue-shaded region (zero) indicate that either both dilutions or the 5-fold dilution resulted in non-detection of the target, and the test was inconclusive.

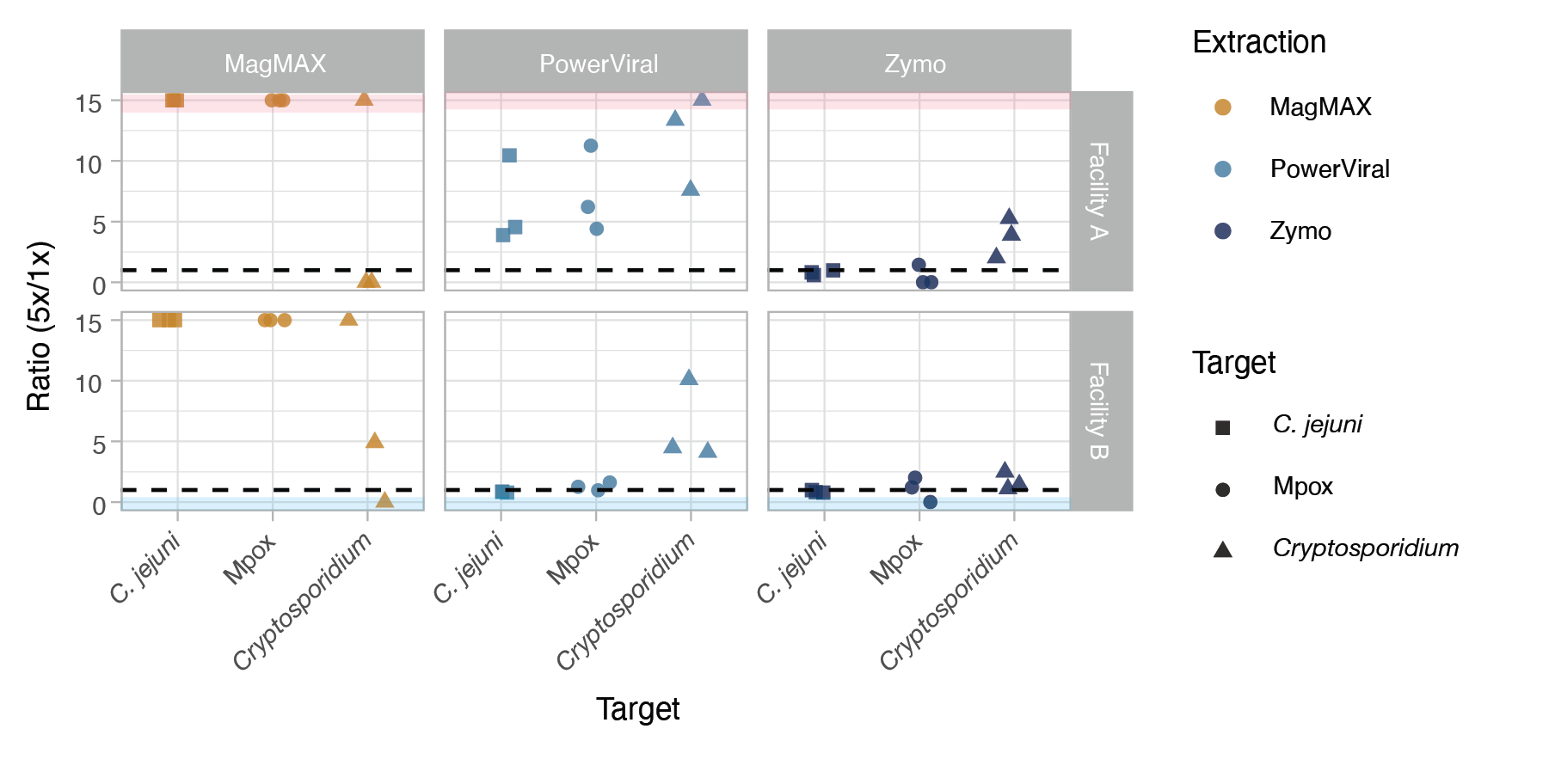

**Figure S3**. dPCR inhibition across three extraction kits for spiked solid samples from two facilities. Samples are considered inhibited if the ratio is greater than 1 (dashed line). Points located at the top of the plot in the red-shaded region (infinity) indicate that the sample was inhibited and the target was detected only in the 5-fold dilution but not in the undiluted template. Conversely, points at the bottom of the plot in the blue-shaded region (zero) indicate that either both dilutions or the 5-fold dilution resulted in non-detection of the target, and the test was inconclusive.

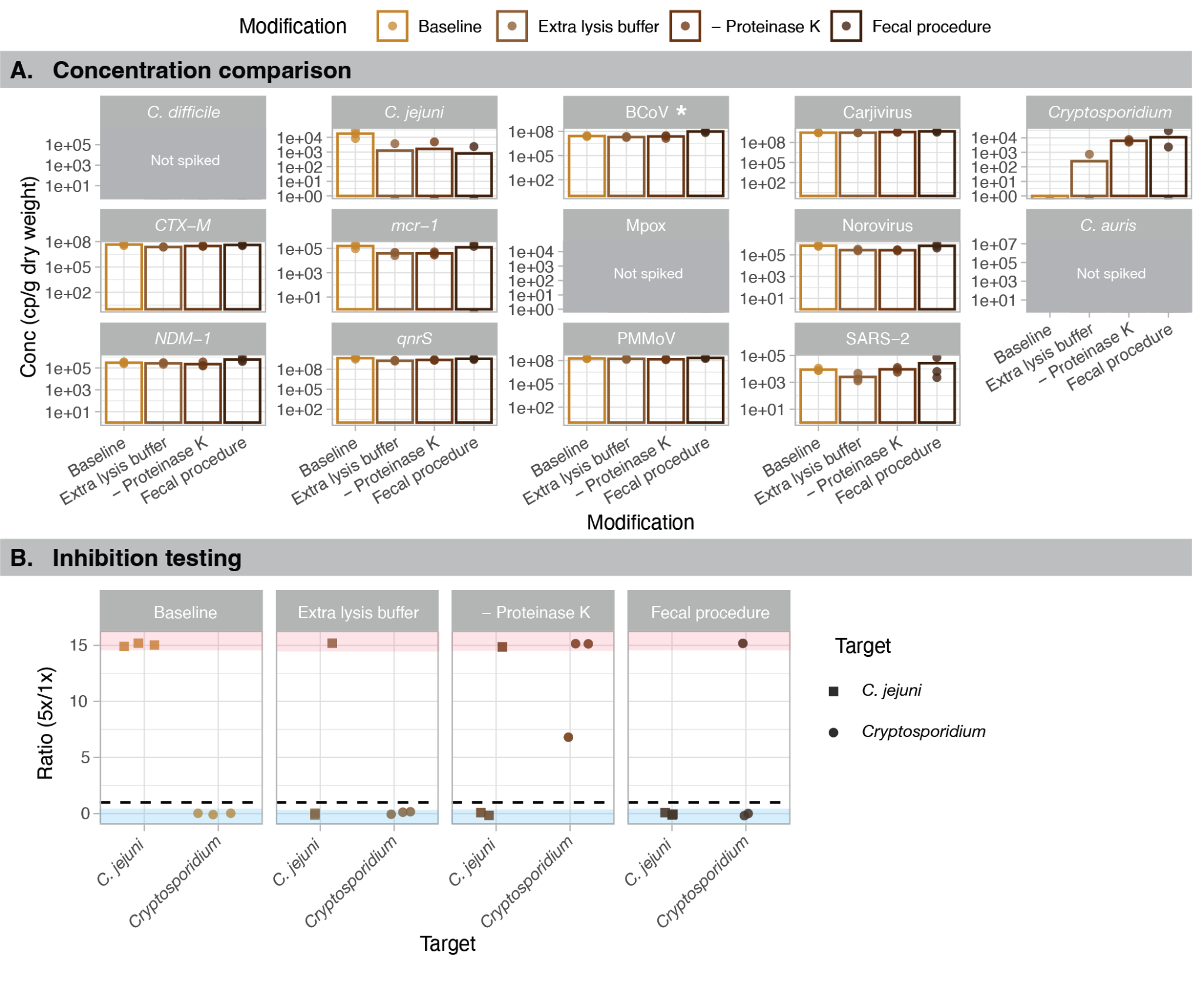

**Figure S4**. **A)** Comparison of MagMAX Microbiome Ultra Nucleic Acid Isolation Kit (baseline) with three modifications geared to alleviate inhibition with addition of extra lysis buffer, omission of proteinase K, or using the fecal procedure, which omits proteinase K but also uses less input material. Each data point is a technical replicate (n = 3) and the bar shows the average concentration. When undetected, the data point shows up under the bar graph. Note that y-axes differ by target and are log-scaled. *C. difficile*, Mpox, and *C. auris* were not spiked and therefore not detected. *Spiked-in targets are indicated by an asterisk. **B)** Inhibition testing results for each modification of MagMAX aimed at alleviating inhibition. Samples are considered inhibited if the ratio is greater than 1 (dashed line). Points located at the top of the plot in the red-shaded region (infinity) indicate that the sample was inhibited and the target was detected only in the 5-fold dilution but not in the undiluted template. Conversely, points at the bottom of the plot in the blue-shaded region (zero) indicate that either both dilutions or the 5-fold dilution resulted in non-detection of the target, and the test was inconclusive.

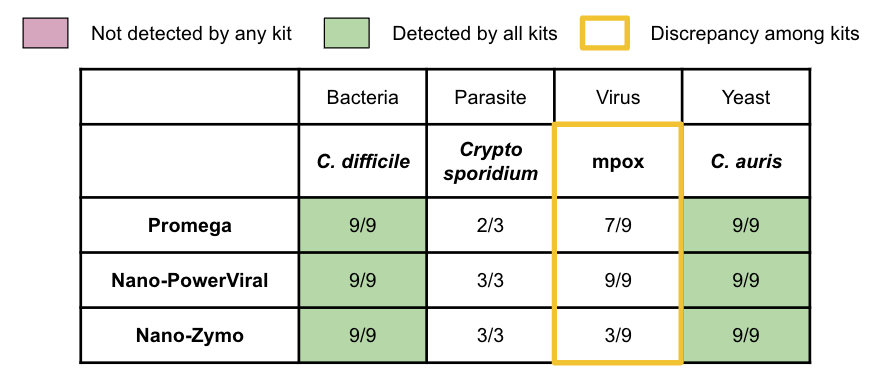

**Figure S5.** **Influent** methods comparison of **spiked-in** targets. The detection rate is shown as the number of total replicates resulting with measurable concentration across three samples used for method comparison (n = 3 for each), except Cryptosporidium, which was only spiked into one sample (n = 3). Color indicates detection agreement level and yellow highlight shows targets with discrepancy between kits in greater than 50% of the replicates.

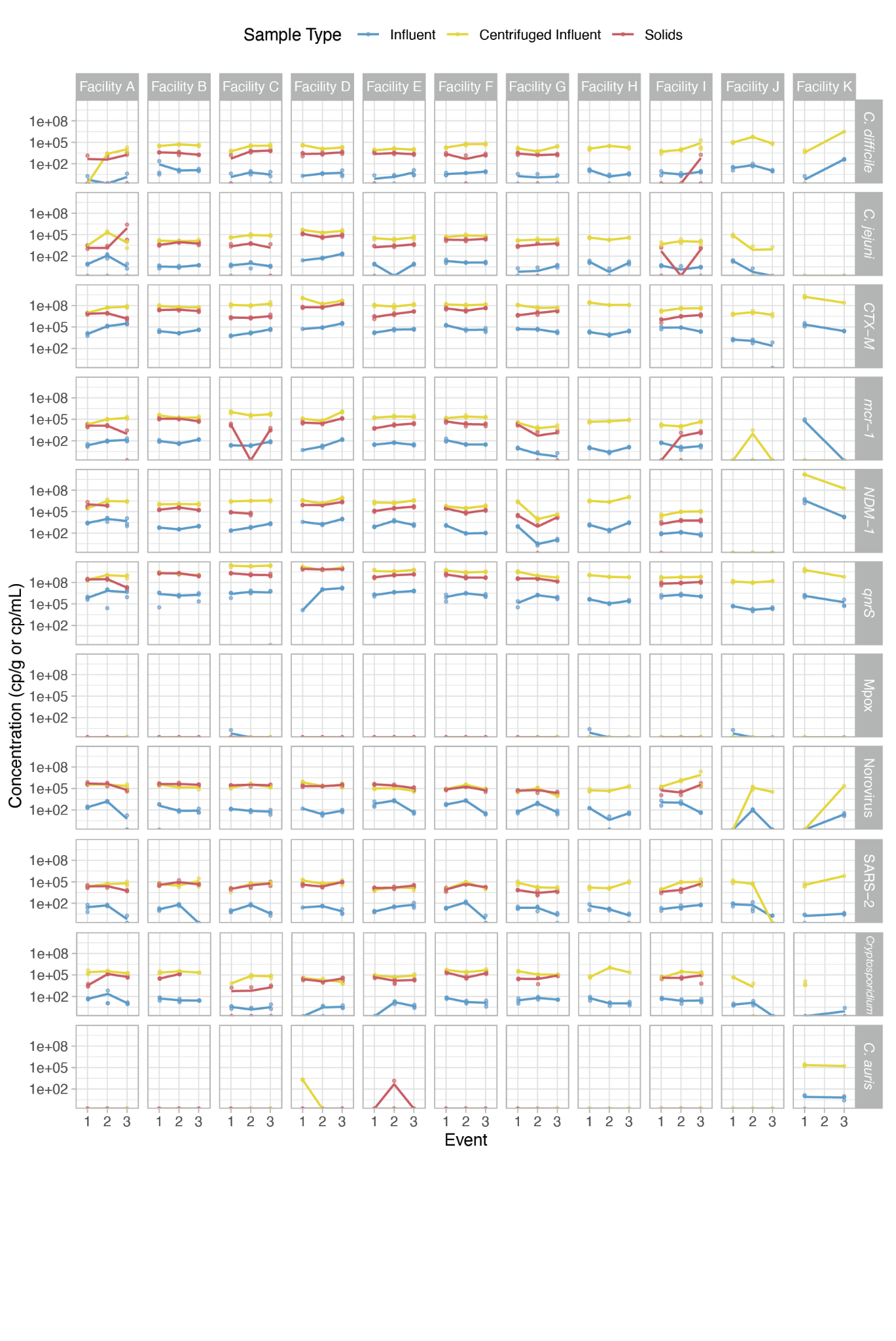

**Figure S6**. Concentration of all samples used in this study in mass basis (cp/g dry weight or c/mL wastewater). Each data point is a replicate and the line connects the average of the replicates across the three sampling events. Note that the y-axis is in log-scale.

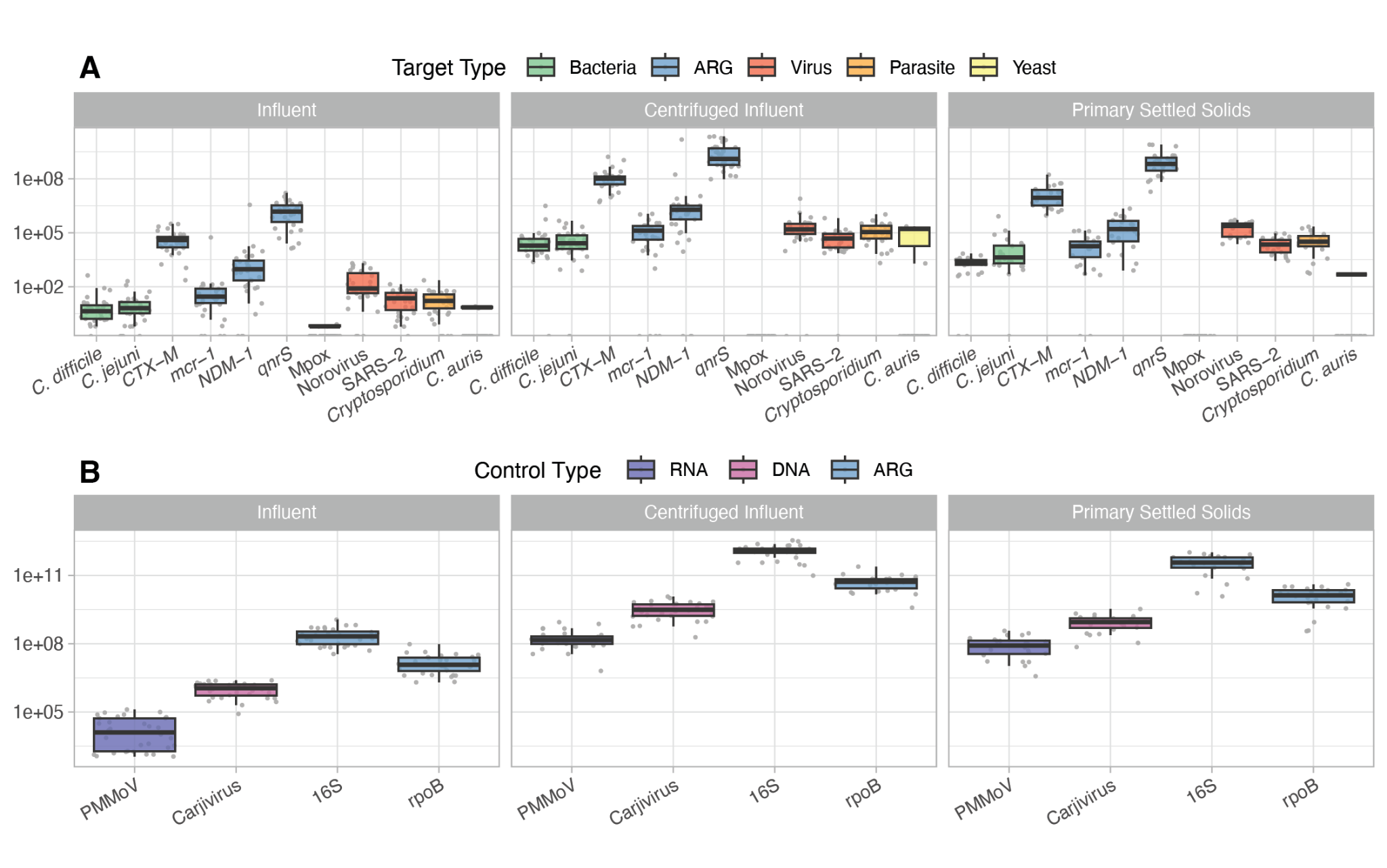

**Figure S7.** Distribution of **A**) targets and **B**) controls in influent (cp/mL), centrifuged influent (cp/g dry weight), and primary settled solid samples (cp/g dry weight). Each data point is averaged across replicates for all available data in each sample type. Boxplot shows median (middle bar in box), 25th and 75th percentile (end of box), and 1.5 times the IQR (extending bars).

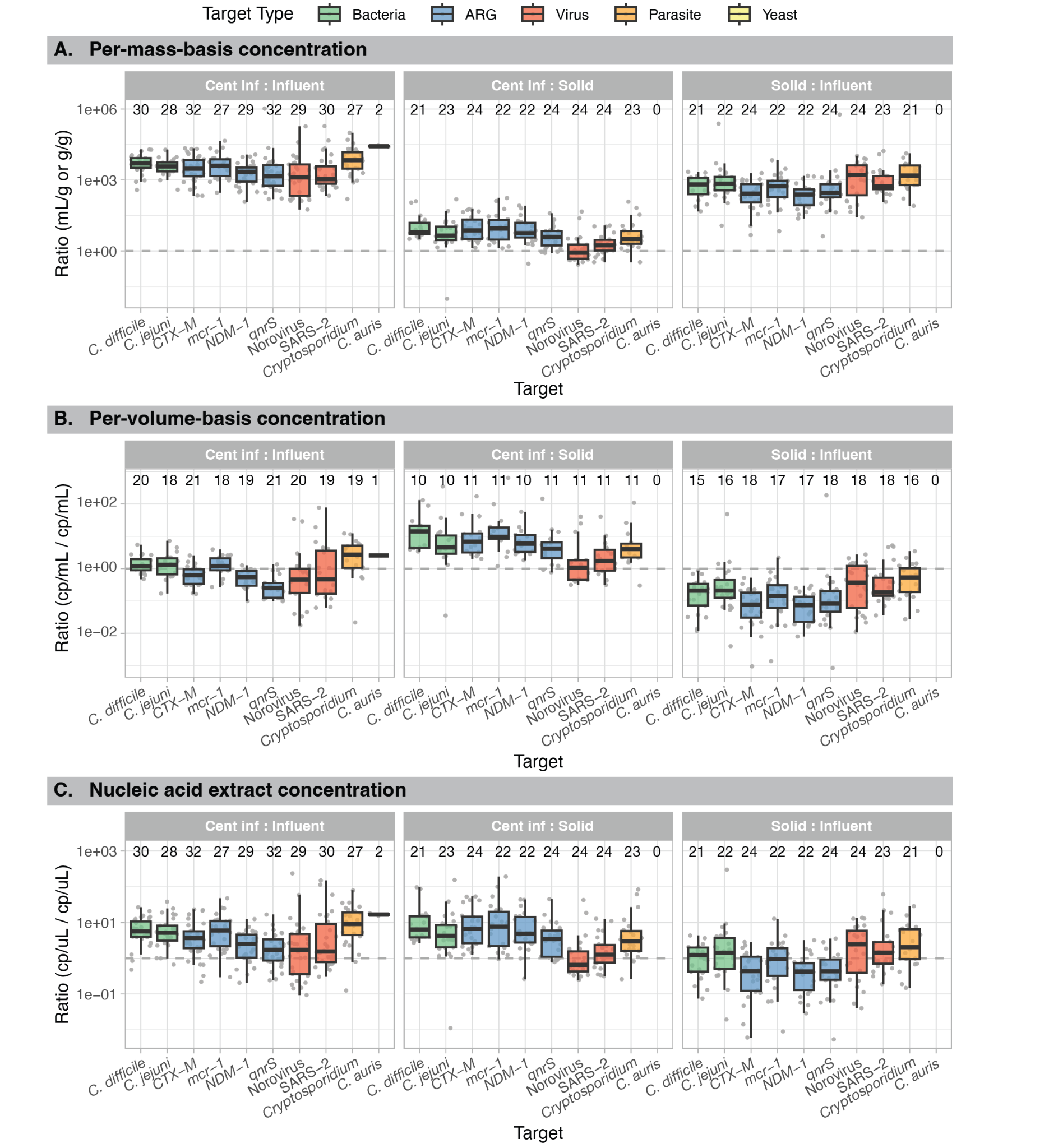

**Figure S8.** Ratio of target concentrations by **A)** per-mass-basis, **B)** extract, and **C)** per-volume-basis in centrifuged influent, whole influent, and primary settled solids. Number of measurements for each target is indicated on top of the plot. Each data point was calculated as a ratio of the target concentration in each date-matched sample. Note log_10_-scale on y-axis. Only data points with detectable concentrations of the target for both matrices are shown. Dashed line represents a ratio of 1.

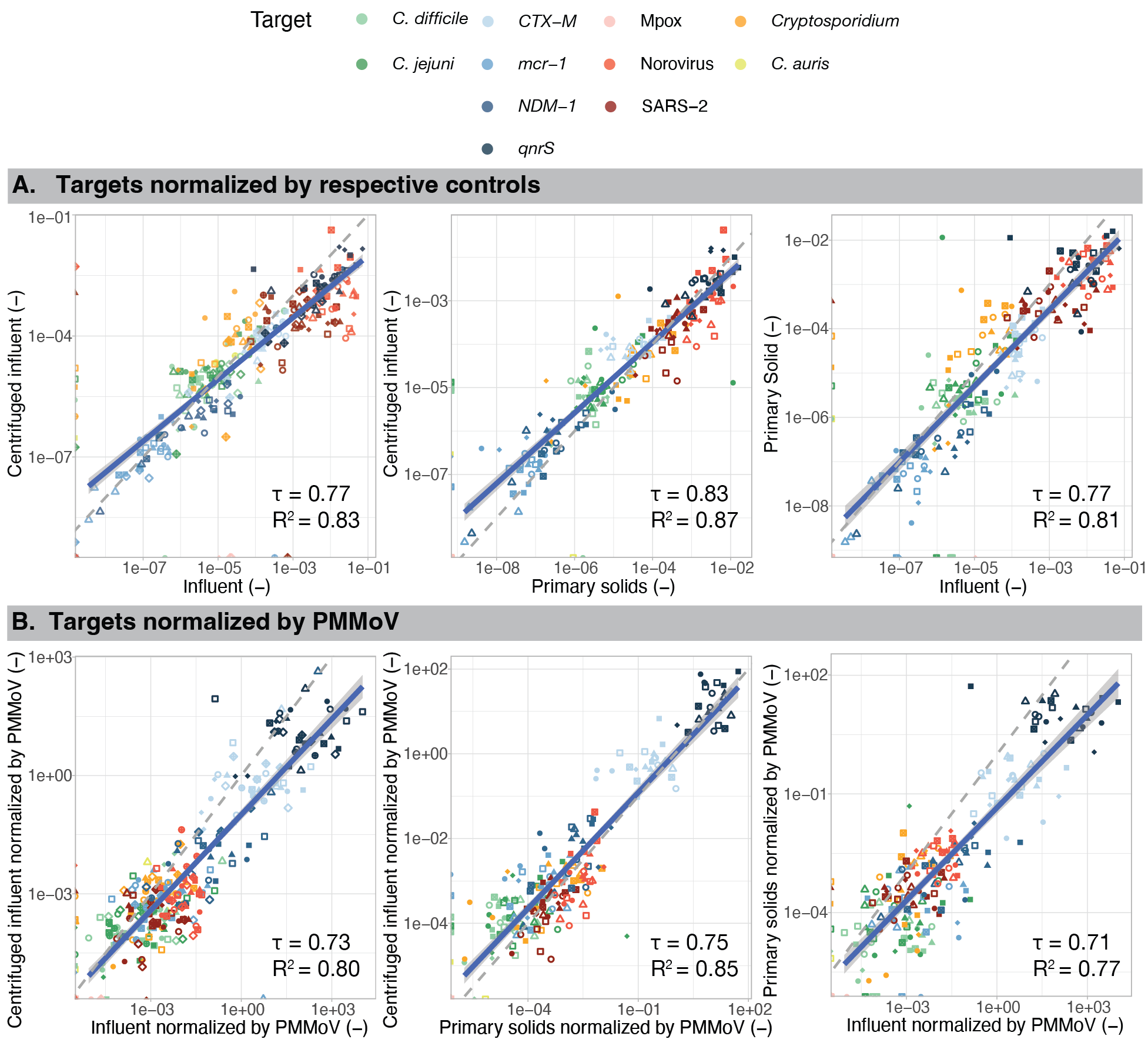

**Figure S9**. Comparison of wastewater concentrations normalized by **A)** respective controls (PMMoV for RNA, carjivirus for DNA, and 16S for ARG) and **B)** PMMoV in paired samples processed by the three sample types. For comparisons with primary settled solids, samples are shown for only the eight wastewater treatment plants that provided paired primary settled solids. Points represent individual targets from one of the three time points at each location. Solid blue line shows linear regression with confidence interval for points that resulted in a measurable concentration, while the gray dashed line shows 1:1 relationship. Note log_10_-scale on both axes. Correlation coefficient Kendall’s tau and R^2^ of the linear regressions shown for each plot.

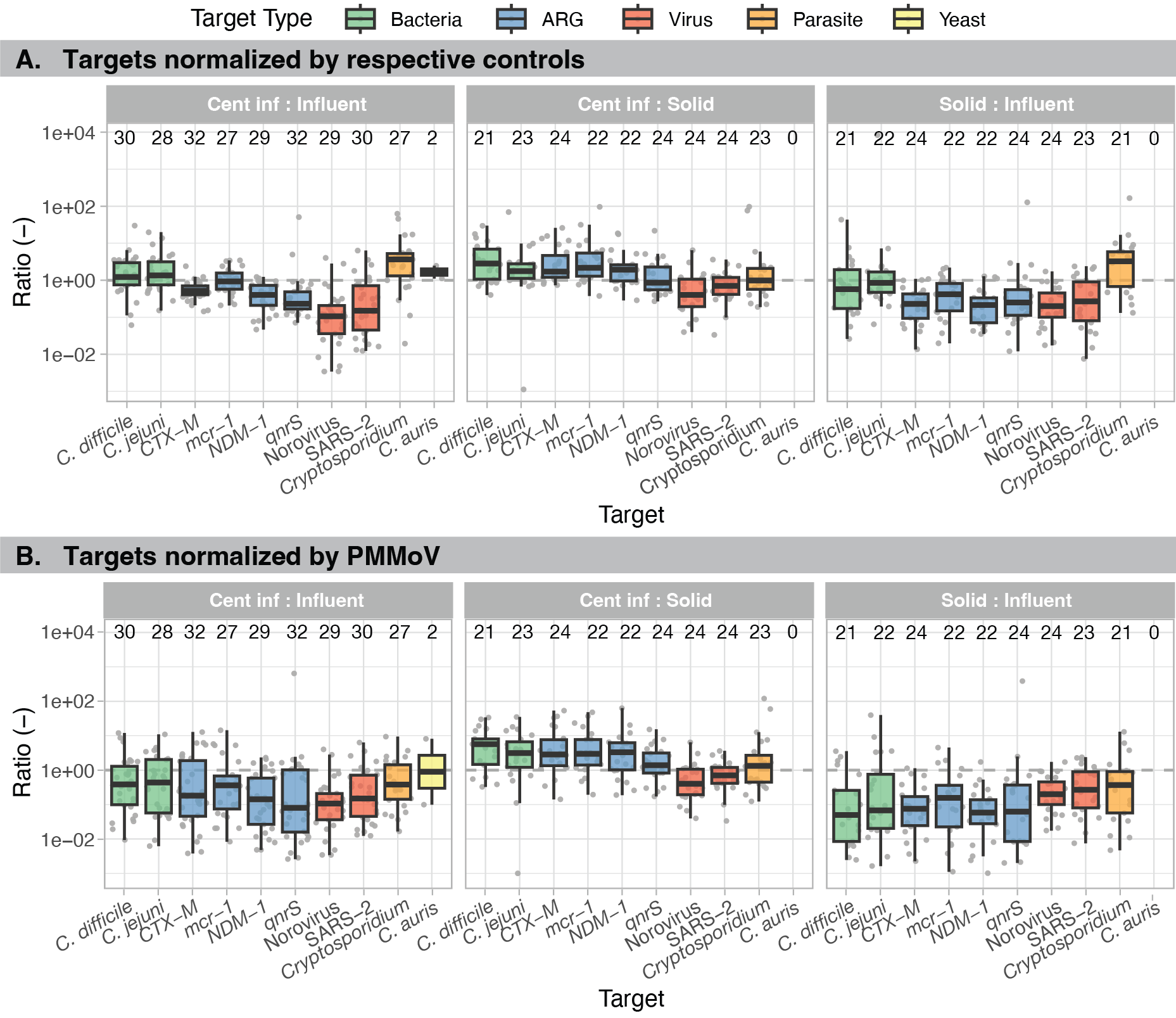

**Figure S10.** Ratio of target concentrations in centrifuged influent, whole influent, and primary settled solids normalized by **A)** respective controls (PMMoV for RNA, carjivirus for DNA, and 16S for ARG) and **B)** PMMoV. Number of measurements for each target is indicated on top of the plot. Note log_10_-scale on y-axis. Only data points with detectable concentrations of the target for both matrices are shown. Dashed line represents a ratio of 1.

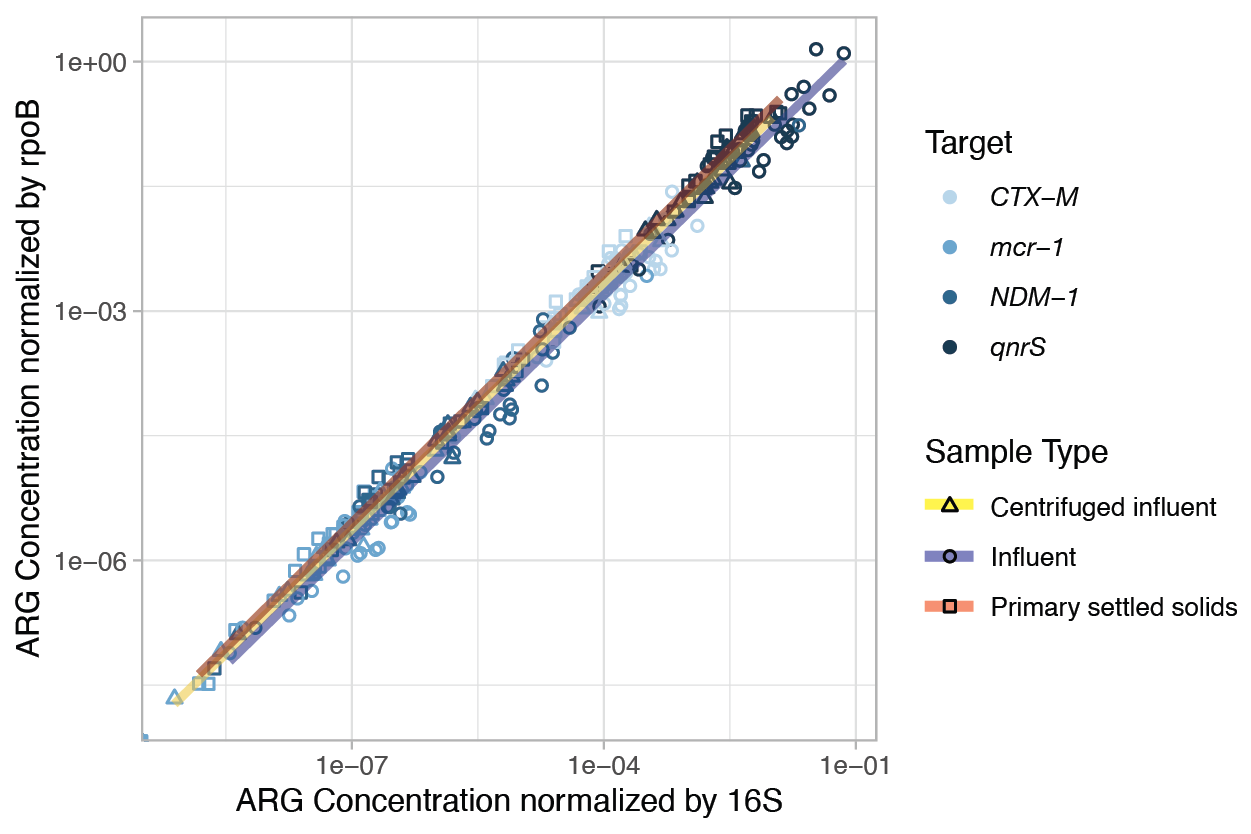

**Figure S11.** Comparison of antibiotic resistance genes (ARGs) normalized by 16S rRNA and rpoB. Linear regression lines that go through each one of the sample types shown in yellow, blue, red for centrifuged influent, influent, and sludge respectively. Note that both axes are in log_10_-scale. The digital PCR assay used here for rpoB resulted in noisy fluorescence signals (known as “rain”) potentially since these assays were designed to capture full diversity of rpoB genes.

### References

1.	Decaro, N. *et al.* Detection of bovine coronavirus using a TaqMan-based real-time RT-PCR assay. *J. Virol. Methods* **151**, 167–171 (2008).

2.	Stachler, E. *et al.* Quantitative CrAssphage PCR Assays for Human Fecal Pollution Measurement. *Environ. Sci. Technol.* **51**, 9146–9154 (2017).

3.	Haramoto, E. *et al.* Occurrence of Pepper Mild Mottle Virus in Drinking Water Sources in Japan. *Appl. Environ. Microbiol.* **79**, 7413–7418 (2013).

4.	He, Y. *et al.* Simultaneous Detection and Differentiation of Campylobacter jejuni, C. coli, and C. lari in Chickens Using a Multiplex Real-Time PCR Assay. *Food Anal. Methods* **3**, 321–329 (2010).

5.	Leach, L., Zhu, Y. & Chaturvedi, S. Development and Validation of a Real-Time PCR Assay for Rapid Detection of Candida auris from Surveillance Samples. *J. Clin. Microbiol.* **56**, e01223-17 (2018).

6.	Shams, A. M., Rose, L. J. & Noble-Wang, J. A. Development of a rapid-viability PCR method for detection of Clostridioides difficile spores from environmental samples. *Anaerobe* **61**, 102077 (2020).

7.	Jothikumar, N., Da Silva, A. J., Moura, I., Qvarnstrom, Y. & Hill, V. R. Detection and differentiation of Cryptosporidium hominis and Cryptosporidium parvum by dual TaqMan assays. *J. Med. Microbiol.* **57**, 1099–1105 (2008).

8.	Li, Y., Olson, V. A., Laue, T., Laker, M. T. & Damon, I. K. Detection of monkeypox virus with real-time PCR assays. *J. Clin. Virol.* **36**, 194–203 (2006).

9.	Fout, G. S. *et al.* Method 1615 Measurement of Enterovirus and Norovirus Occurrence in Water by Culture and RT-qPCR. (2014).

10.	Pholwat, S. *et al.* Genotypic antimicrobial resistance assays for use on E. coli isolates and stool specimens. *PLOS ONE* **14**, e0216747 (2019).

11.	Chavda, K. D. *et al.* Evaluation of a Multiplex PCR Assay To Rapidly Detect Enterobacteriaceae with a Broad Range of β-Lactamases Directly from Perianal Swabs. *Antimicrob. Agents Chemother.* **60**, 6957–6961 (2016).

12.	Suzuki, M. T., Taylor, L. T. & DeLong, E. F. Quantitative Analysis of Small-Subunit rRNA Genes in Mixed Microbial Populations via 5′-Nuclease Assays. *Appl. Environ. Microbiol.* **66**, 4605–4614 (2000).

13.	Takahashi, H., Konuma, H. & Hara-Kudo, Y. Development of a Quantitative Real-Time PCR Method To Enumerate Total Bacterial Counts in Ready-to-Eat Fruits and Vegetables. *J. Food Prot.* **69**, 2504–2508 (2006).
